# Effects of Non-Cardiac/Non-neurologic Surgery and Anesthesia on the CSF Proteome, and Modulation by the APOE Mimetic Peptide CN-105

**DOI:** 10.1101/2025.10.06.25337433

**Authors:** Haotian Zou, Michael W. Lutz, Matthew W. Foster, Mary Cooter Wright, Marlene J. Violette, Sheng Luo, Daniel Laskowitz, Michael J. Devinney, Miles Berger, the MARBLE Teams INTUIT & MADCO-PC Study

**Affiliations:** Department of Biostatistics & Bioinformatics, Duke University School of Medicine, Durham, NC, USA; Department of Neurology, Duke University School of Medicine, Durham, NC, USA; Duke-UNC Alzheimers Disease Research Center, Duke University Medical Center, Durham, NC, USA; Proteomics and Metabolomics Core Facility, Duke University School of Medicine, Durham, NC, USA; Department of Medicine, Duke University School of Medicine, Durham, NC, USA; Department of Anesthesiology, Duke University School of Medicine, Durham, NC, USA; Department of Anesthesiology, Perioperative and Pain Medicine, Stanford University School of Medicine & Stanford Health, Stanford CA, USA

**Keywords:** Surgery, CSF, Proteome, APOE, CN-105

## Abstract

**Background:** Neuroinflammation is thought to contribute to postoperative central nervous system (CNS)-related complications, and the apolipoprotein E (APOE)-mimetic peptide CN-105 blocks neuroinflammation in animal models. Thus, here we examined postoperative cerebrospinal fluid (CSF) proteome changes and their modulation by CN-105.

**Objective:** To evaluate postoperative changes in the CSF proteome, and their modulation by the APOE mimetic peptide CN-105.

**Methods:** We performed mass spectrometry-based proteomics on preoperative and 24-hour postoperative CSF samples from 137 non-cardiac/non-neurologic surgery MARBLE trial patients (age>60; ct.gov identifier: NCT03802396), who were randomized to receive APOE mimetic peptide CN-105 (or placebo), and in an independent replication cohort. Linear regression evaluated postoperative changes in CSF protein and pathway scores (quantified by singe set gene set enrichment assay [ssGSEA] pathway scores), with false discovery rate (FDR)-based multiple comparison correction.

**Results:** 24-hour postoperative changes were observed in 881 of 2,086 proteins (57 of which showed a log2 fold change >0.5 or <-0.5) and in 1001 of 1854 pathways (p-FDR<0.05). Similar magnitude temporal effects were seen in these proteins and pathways in a replication cohort. The most significantly upregulated CSF pathways involved smooth muscle cell migration/regulation or apoptotic signaling/regulation; the 5 most significantly downregulated CSF pathways included NF-κB signal transduction regulation, leukocyte apoptotic process, and sulfur and proteoglycan metabolic processes. There was no significant CN-105 effect on 24-hour postoperative changes in CSF protein levels or ssGSEA pathway scores (p-FDR>0.05 for all).

**Conclusions:** Significant postoperative changes occurred in over 40% of proteins and over 50% of pathways in the CSF, particularly in smooth muscle, endothelial, leukocyte, and apoptosis pathways.

## BACKGROUND

Perioperative care can lead to postoperative neurocognitive complications ranging from postoperative delirium to postoperative neurocognitive disorder and stroke.^1–3^ The underlying molecular basis for these disorders is unclear, though inflammation has been postulated to play a key role; this can be examined using pre- and post-operative cerebrospinal fluid (CSF) samples.^4^ Indeed, CSF mass spectrometry studies have identified specific protein pathways implicated in Alzheimer’s Disease,^5–7^ and a pilot mass spectrometry study identified differences in CSF complement pathway levels over time among patients with postoperative cognitive dysfunction.^5^ Yet, few if any studies have systematically studied postoperative CSF proteome changes among a large cohort of older non-cardiac/non-neurologic surgery patients, which would be a first step towards understanding what postoperative central nervous system (CNS) proteomic changes normally occur, and which might be dysregulated in patients with perioperative neurocognitive disorders.

Additionally, there are clearly some patients who have worse long term cognitive trajectories after surgery, such as those who carry the E4 allele of apolipoprotein E (ie *APOE4*). *APOE4* carriers (versus non-carriers) have worse long term cognitive trajectories in the 5 years following surgery,^8^ and *APOE4* carriers who underwent surgery had worse long term cognitive decline than non-*APOE4* carriers who underwent surgery, and as compared to both *APOE4* carriers and non-*APOE4* carriers who did not undergo surgery.^9^ *APOE4* carriers have altered CSF levels of inflammation-related proteins such as C-reactive protein (CRP), complement and YKL-40/CH3L1,^10, 11^ which may contribute to long term neurocognitive decline. Further, modulating APOE protein signaling with the APOE mimetic penta-peptide CN-105 reduces neuro-inflammation and improved outcomes in mice after multiple types of brain injury.^12, 13^ CN-105 was safely administered to humans in a phase I trial,^14^ and to older surgical patients in a phase II trial.^15^ Thus, here we addressed two questions. First, what are the changes in the CSF proteome from before to 24 hours after a variety of non-cardiac/non-neurologic surgeries in older adults (ie age ≥ 60 years)? Second, does modulating APOE protein signaling with CN-105 in older patients alter these postoperative CSF protein and/or pathway changes?

## METHODS

### Study Overview and Intervention

We analyzed CSF samples from the triple-blind randomized phase II trial Modulating APOE to Reduce Brain infLammation and postoperative dElirium (MARBLE), which aimed to evaluate the safety and feasibility of the investigational product CN-105 for preventing delirium in older adults (age ≥ 60 years) after non-cardiac/neurologic surgery (see Supplemental Digital Content Section 1 for additional MARBLE study information). MARBLE was approved by the Western Institutional Review Board and registered with clinicaltrials.gov prior to study initiation (ct.gov identifier: NCT03802396). MARBLE excluded prisoners, those scheduled to receive chemotherapy with detrimental cognitive effects within 6 weeks following surgery, those unable to undergo a lumbar puncture (due to anticoagulation, severe anxiety, or other contraindication) or inappropriate for inclusion based on principal investigator judgement. The primary and secondary outcomes of the MARBLE trial have been published separately.^15^

MARBLE participants were randomized to receive either placebo, 0.1 mg/kg, 0.5 mg/kg, or 1 mg/kg of CN-105 every 6 hours, starting just before surgery until 3 days later (for 13 doses maximum). After enrollment, MARBLE patients underwent lumbar punctures for cerebrospinal fluid (CSF) collection (as described^4^) before receiving the first dose of CN-105 or placebo, and again 24 hours after the start of surgery.

### Proteomics Data and Study Population

We performed mass spectrometry-based proteomics on CSF samples from all 137 MARBLE participants who underwent preoperative and 24 hour postoperative lumbar punctures; see Supplemental Digital Content Section 1 for details on CSF proteomic data acquisition and processing. Protein intensities were log2 transformed for normality.

### Replication Cohort

To replicate/validate our findings from the MARBLE study CSF samples in an independent cohort, we also analyzed mass spectrometry data from CSF samples obtained before and 24 hours after surgery among 96 patients from the MADCO-PC and INTUIT studies (see Supplemental Digital Content Section 3- Replication Cohort Description for details).^16, 17^ MADCO-PC and INTUIT were approved by the Duke IRB, and registered with clinicaltrials.gov (NCT01993836, NCT03273335). For validation, we selected the CSF proteins or pathways that showed a false discovery rate (FDR) significant temporal effect in the MARBLE cohort. Then we analyzed the temporal effects of surgery on these protein or pathway levels in this replication cohort via the same methods as in the MARBLE cohort and reported the nominal p-values.

### Statistical Analysis

#### Missing Data Imputation and Temporal Effects on Protein Intensity and Pathway Enrichment Scores

We imputed missing protein intensities via the random forest method (see Supplemental Digital Content Section 1), and then examined changes in each CSF protein from the pre-operative to 24-hour postoperative timepoint via univariable linear regression models, with multiple comparison correction using false discovery rate (FDR) across the p-values from the 2,086 CSF proteins, with α=0.05^18^. To rule out the possibility of confounding by baseline patient characteristics or surgery type, we also examined changes in each CSF protein and pathway over this time interval using multivariable adjusted linear regression models.

To analyze the temporal effect of non-cardiac/neurologic surgery on protein pathway enrichment scores, we performed single-sample gene set enrichment analysis (ssGSEA) to provide a pathway enrichment score for each pathway in each CSF sample.^19, 20^ We used the Gene Ontology database for biological processes (GO-BP), version 2023.2, and extracted ssGSEA scores from 1854 pathways. We computed the difference between the raw ssGSEA scores at the preoperative and 24 hour postoperative timepoints for each pathway, and modeled these differences via both independent univariable linear regression models and independent multivariable adjusted linear regression models, with FDR adjustment across the 1,854 GO-BP pathways in the analysis.

#### APOE4 effect on Pre to Postoperative Changes in CSF Protein

In a sensitivity analysis, we examine the effect of *APOE4* carrier status on pre to 24 hr postoperative CSF protein changes via independent linear mixed models with CSF protein levels (pre-operative and 24-hour post-operative) as the dependent variable, for each CSF protein. We included the following covariates: timepoint (pre-operative or 24-hour post-operative), *APOE4* carrier status, interaction between timepoint and *APOE4* carrier status, and a random intercept. We tested the effects of time and *APOE4* carrier status, and their interaction effect.

#### Unbiased Analysis of CN-105 Effect on Changes in Protein Intensity and Pathway Enrichment Score

We assessed the effect of CN-105 on pre-operative to 24-hour post-operative changes in CSF protein intensity levels and pathway enrichment scores using both univariable and multivariable adjusted linear models, as described above. To assess the effect of CN-105 on pre- to 24-hour post-operative changes in pathway enrichment scores, we first selected pathways that showed a significant change from before to 24 hours after surgery. We fit both univariable and multivariable adjusted linear models for pathway enrichment score temporal change, using a regression coefficient for group (CN-105 vs placebo).

## RESULTS

### Baseline Characteristics, and Temporal Effects on Protein Intensity and Pathway Enrichment Score in the MARBLE cohort

Table 1 displays the baseline MARBLE cohort characteristics; see Supplemental Digital Content Figure S1 for CONSORT diagram, and Supplemental Digital Content Section 1 for the missing data imputation approach and performance. Of 2,086 proteins with a missingness of ≤ 80% at both time points in our MARBLE study mass spectrometry dataset, after FDR adjustment, 881 proteins (42.2%) showed a significant intensity level change from before to 24 hours after non-cardiac/neurologic surgery (Fig 1A for the volcano plot), of which 57 (6.4%) proteins showed a log_2_ fold change above 0.5 or below -0.5 (i.e., these 57 proteins showed a moderate to large effect size in the log_2_ fold change).^21^ Table 2 displays the most significantly up-regulated proteins and down-regulated proteins with log_2_ FC above 0.5 or below -0.5, from univariable linear models that assessed protein level changes from before to 24 hours after surgery. The 5 most significantly changed protein levels (by t values) after surgery were FSTL3 (Follistatin Like 3; upregulated), SDF1 (Stromal Cell-Derived Factor 1; downregulated), IBP3 (Insulin-like growth factor-binding protein 3; upregulated), SRGN (Serglycin; upregulated), and SAA1 (Serum amyloid A-1 protein; upregulated). All of these top 5 postoperative protein level changes remained highly significant (with the same directionality) in an FDR-corrected multivariable analysis adjusted for numerous baseline patient characteristics (Supplemental Digital Content Table S7, Supplemental Digital Content Data Sheet 6), which rules out the possibility that these postoperative protein level changes were confounded by or dependent on specific patient characteristics. The magnitude of these postoperative CSF protein level changes were also similar across patients who underwent different types of surgery (Supplemental Digital Content Table S1).

**Figure 1.**
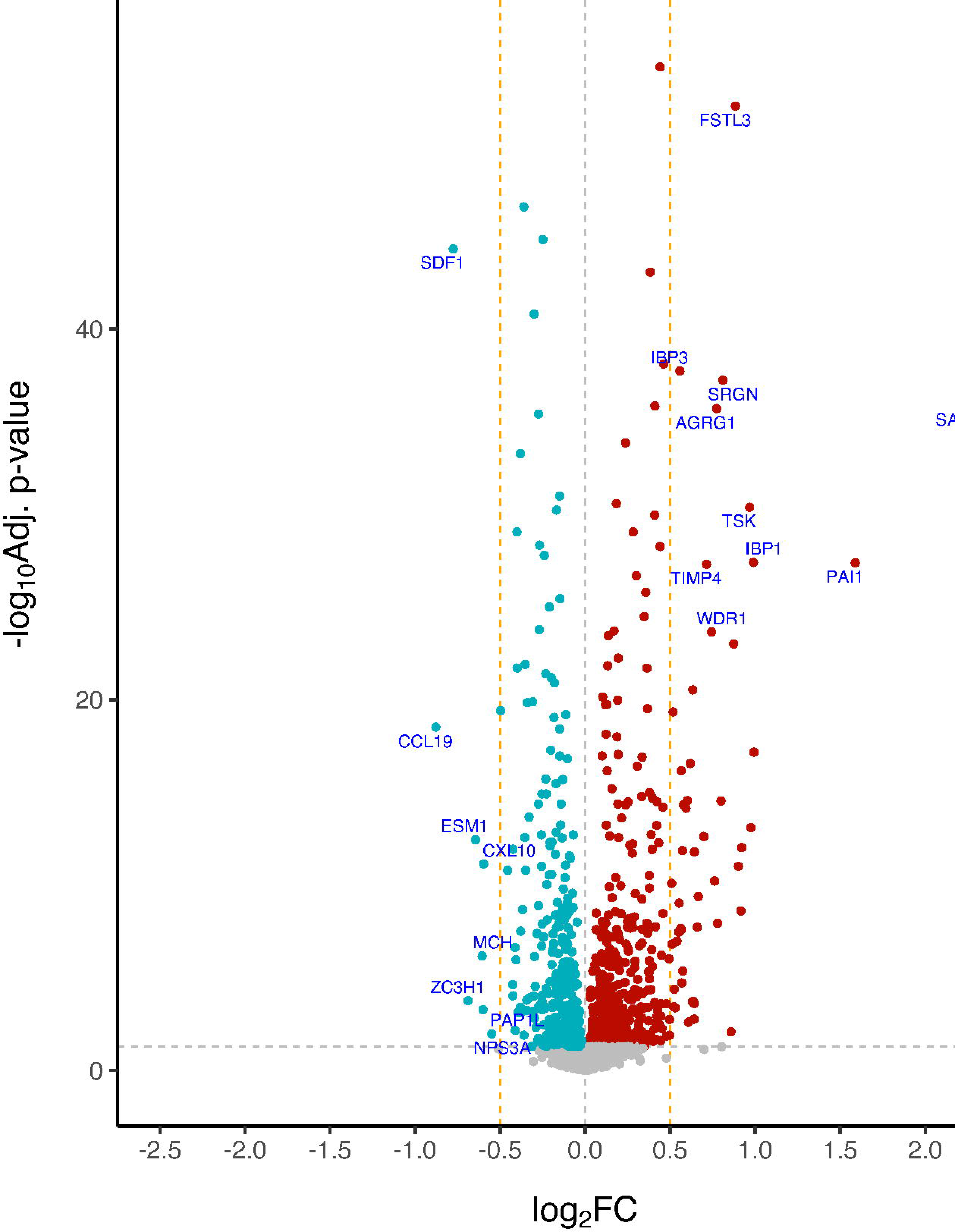
A) Volcano plot for the effect of time on CSF protein levels in the MARBLE cohort. The x-axis is the log_2_ fold change of the univariate differences in protein intensity levels from before to 24 hours after surgery. The y-axis is the -log_10_ base adjusted p-value. The horizontal dashed line indicates -log_10_ base adjusted p-value of 0.05. The vertical dashed lines indicate log_2_ fold change threshold of 0.5 and -0.5 (i.e., these proteins show moderate to large effects). The 10 up-regulated proteins and 8 down-regulated proteins sorted by p-values and log_2_ fold change threshold above 0.5 or below -0.5 are labelled below. B) Volcano Plot for the univariate effect of time on CSF protein pathways in the MARBLE cohort, using the GO database for biological processes (GO-BP). The horizontal dashed line indicates -log_10_ base adjusted p-value of 0.05. The pathways with top 10 p-values are labelled below.

**Table 1.**
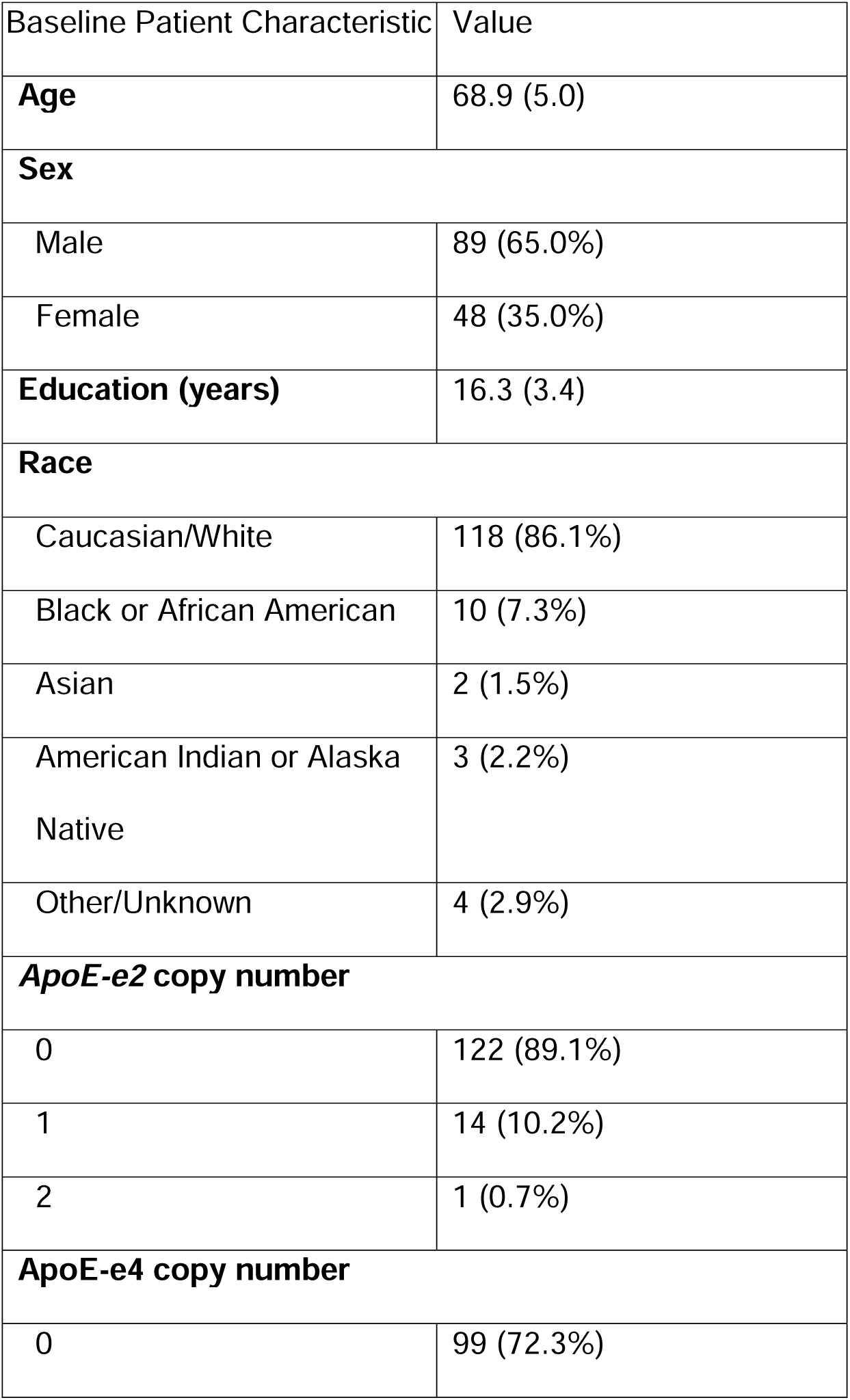
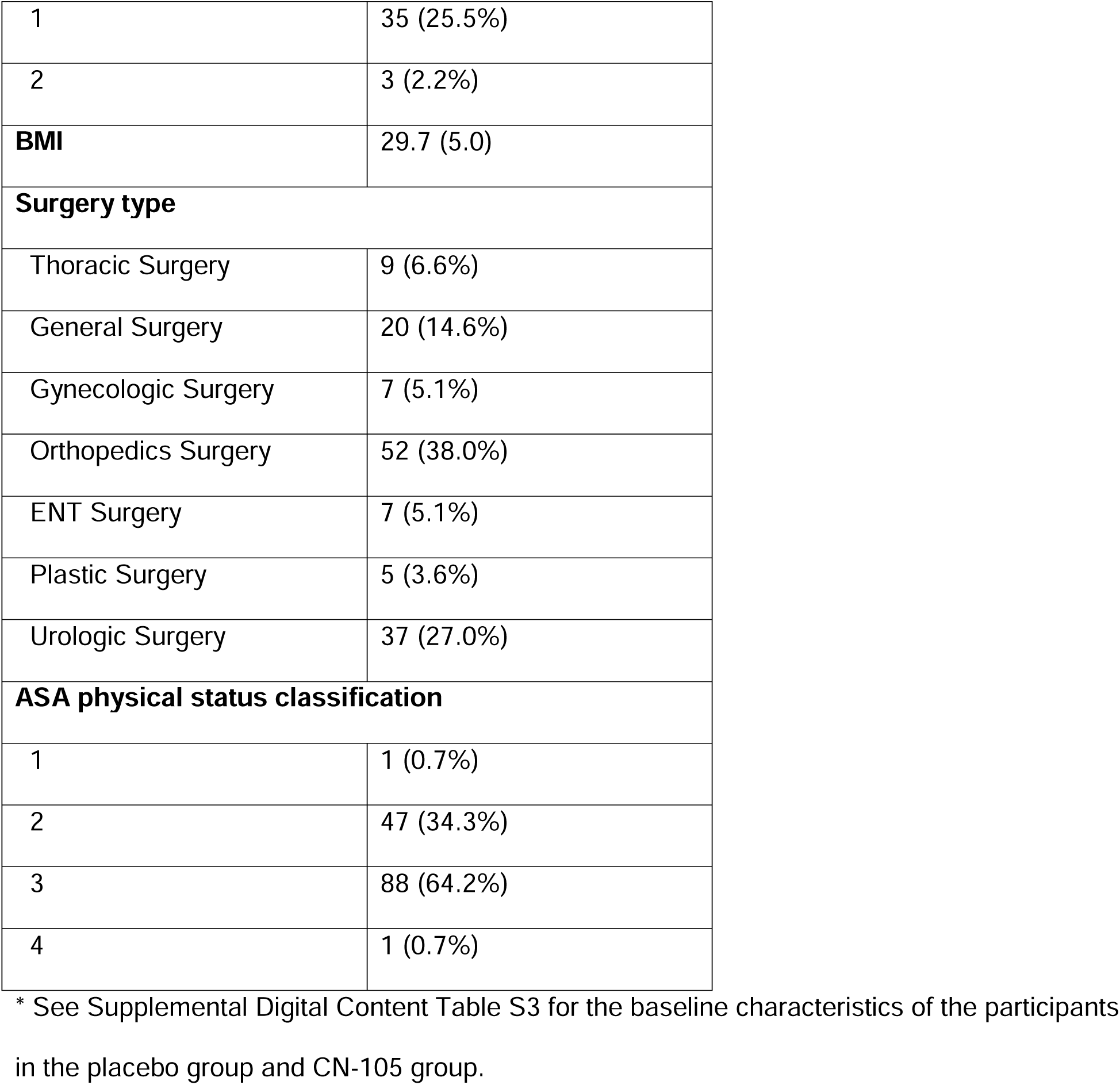
Baseline characteristics of the 137* MARBLE study participants with preoperative and 24 hr postoperative CSF samples. The numbers are presented as mean (sd) or count (percentage).

**Table 2.** The 10 up-regulated and 8 down-regulated proteins (sorted by p-value) from univariable linear models (t-tests) that assessed changes in CSF protein levels from before to 24 hours after surgery in the MARBLE patient cohort, with a log2 fold change above 0.5 or below -0.5 (i.e., these proteins show moderate to large effects). Positive fold change values (Log_2_FC) indicate an increase in the given protein level from before to 24 hours after surgery, and *vice versa*.

| **Up-regulated proteins** |  | **Log<sub>2</sub> FC** | **Std.Err** | **95% CI** | **t** | **p-value** | **Adj.P** |
| --- | --- | --- | --- | --- | --- | --- | --- |
| FSTL3 | Follistatin-related protein 3 | 0.884 | 0.033 | (0.819, 0.949) | 26.608 | 9.14E-56 | 9.54E-53 |
| IBP3 | Insulin-like growth factor-binding protein 3 | 0.557 | 0.029 | (0.500, 0.614) | 19.323 | 8.00E-41 | 1.85E-38 |
| SRGN | Serglycin | 0.809 | 0.042 | (0.727, 0.891) | 19.081 | 2.80E-40 | 5.85E-38 |
| SAA1 | Serum amyloid A-1 protein | 2.255 | 0.122 | (2.016, 2.494) | 18.436 | 8.27E-39 | 1.44E-36 |
| AGRG1 | Adhesion G-protein coupled receptor G1 | 0.773 | 0.042 | (0.691, 0.855) | 18.361 | 1.23E-38 | 1.97E-36 |
| TSK | Tsukushin | 0.968 | 0.060 | (0.850, 1.086) | 16.036 | 3.77E-33 | 4.14E-31 |
| IBP1 | Insulin-like growth factor-binding protein 1 | 0.990 | 0.067 | (0.859, 1.121) | 14.756 | 5.09E-30 | 3.94E-28 |
| PAI1 | Plasminogen activator inhibitor 1 | 1.589 | 0.108 | (1.377, 1.801) | 14.741 | 5.56E-30 | 4.15E-28 |
| TIMP4 | Metalloproteinase inhibitor 4 | 0.714 | 0.049 | (0.618, 0.810) | 14.705 | 6.83E-30 | 4.91E-28 |
| WDR1 | WD repeat-containing protein 1 | 0.743 | 0.056 | (0.633, 0.853) | 13.204 | 3.91E-26 | 2.20E-24 |
| **Down-regulated proteins** |  | **Log<sub>2</sub> FC** | **Std.Err** | **95% CI** | **T** | **p-value** | **Adj.P** |
| SDF1 | Stromal cell-derived factor 1 | -0.776 | 0.034 | (-0.843, -0.709) | -22.489 | 1.16E-47 | 4.85E-45 |
| CCL19 | C-C motif chemokine 19 | -0.878 | 0.079 | (-1.033, -0.723) | -11.104 | 8.63E-21 | 3.00E-19 |
| ESM1 | Endothelial cell-specific molecule 1 | -0.645 | 0.075 | (-0.792, -0.498) | -8.580 | 1.90E-14 | 3.61E-13 |
| CXL10 | C-X-C motif chemokine 10 | -0.597 | 0.074 | (-0.742, -0.452) | -8.019 | 4.38E-13 | 7.24E-12 |
| MCH | Pro-MCH | -0.606 | 0.107 | (-0.816, -0.396) | -5.680 | 7.83E-08 | 6.66E-07 |
| ZC3H1 | Zinc finger C3H1 domain-containing protein | -0.689 | 0.160 | (-1.003, -0.375) | -4.309 | 3.12E-05 | 1.74E-04 |
| PAP1L | Polyadenylate-binding protein 1-like | -0.601 | 0.151 | (-0.897, -0.305) | -3.989 | 1.08E-04 | 5.24E-04 |
| NPS3A | Protein NipSnap homolog 3A | -0.550 | 0.183 | (-0.909, -0.191) | -2.999 | 3.22E-03 | 1.06E-02 |

\* Abbreviations: Std.Err: standard error; Adj.P: adjusted p-value. FC: fold change. CI: confidence interval.
\* See Supplemental Digital Content Table S7 for multivariable model adjusted for CN-105, age, sex, education years, *ApoE*-e2 copy number, *ApoE*-e4 copy number, BMI, surgery type, ASA physical status classification, and baseline protein intensity.

1001 of 1854 pathways (54.0%) showed a significant change (i.e. p-FDR<0.05) in enrichment scores from before to 24 hours after surgery in the MARBLE dataset (Fig 1B; Table 3). Supplemental Digital Content Tables S2a and S2b list the genes for each pathway that was up- or down-regulated after surgery, respectively. The 5 most significantly upregulated pathways were negative regulation of smooth muscle cell migration, negative regulation of endothelial cell apoptotic process, extrinsic apoptotic signaling pathway via death domain receptors, regulation of extrinsic apoptotic signaling pathway via death domain receptors, and smooth muscle cell migration. The 5 most significantly downregulated pathways were regulation of non-canonical NF κB signal transduction, negative regulation of leukocyte apoptotic process, sulfur compound metabolic process, proteoglycan metabolic process, and leukocyte apoptotic process. All of these top postoperative pathway enrichment score changes (i.e., 10 out of 10 up-regulated pathways, and 10 out of 10 down-regulated pathways) remained highly significant (and in the same direction) in an FDR-corrected multivariable analysis that was adjusted for numerous baseline patient characteristics (Supplemental Digital Content Table S8, Supplemental Digital Content Data Sheet 7), which rules out the possibility that these postoperative pathway enrichment score changes were confounded by or dependent on specific patient characteristics.

**Table 3.** The 10 up-regulated and 10 down-regulated pathways (sorted by p-value) from the univariable linear models that assessed the temporal effect of the pathway enrichment score from before to 24 hours after surgery, based on the Gene Ontology database for biological processes (GO-BP) in the MARBLE cohort. The top panel displays pathways that are the most highly up-regulated; the bottom panel displays pathways that are the most down-regulated. Beta coefficients are from linear models that assessed the effect of time (ie from before to 24 hrs after anesthesia and surgery) on the ssGSEA enrichment scores.

| **Up-regulated pathways** | **Beta** | **Std.Err** | **95% CI** | **t** | **p-value** | **Adj.P** |
| --- | --- | --- | --- | --- | --- | --- |
| negative regulation of smooth muscle cell migration | 77.574 | 4.183 | (69.375, 85.773) | 18.547 | 4.60E-39 | 8.53E-36 |
| negative regulation of endothelial cell apoptotic process | 93.981 | 5.417 | (83.364, 104.598) | 17.349 | 2.76E-36 | 2.56E-33 |
| extrinsic apoptotic signaling pathway via death domain receptors | 67.583 | 4.641 | (58.487, 76.679) | 14.563 | 1.54E-29 | 9.49E-27 |
| regulation of extrinsic apoptotic signaling pathway via death domain receptors | 79.959 | 5.556 | (69.069, 90.849) | 14.392 | 4.08E-29 | 1.89E-26 |
| smooth muscle cell migration | 40.878 | 3.068 | (34.865, 46.891) | 13.323 | 1.96E-26 | 7.28E-24 |
| regulation of fibrinolysis | 43.100 | 3.474 | (36.291, 49.909) | 12.407 | 4.10E-24 | 1.27E-21 |
| muscle cell migration | 35.009 | 2.883 | (29.358, 40.660) | 12.144 | 1.92E-23 | 5.08E-21 |
| endothelial cell apoptotic process | 61.060 | 5.199 | (50.870, 71.250) | 11.744 | 2.01E-22 | 4.65E-20 |
| angiogenesis involved in wound healing | 67.270 | 5.860 | (55.784, 78.756) | 11.480 | 9.48E-22 | 1.95E-19 |
| actin filament organization | 36.949 | 3.245 | (30.589, 43.309) | 11.386 | 1.65E-21 | 3.05E-19 |
| **Down-regulated pathways** | **Beta** | **Std.Err** | **95% CI** | **t** | **p-value** | **Adj.P** |
| regulation of non-canonical NF κB signal transduction | -50.390 | 4.940 | (-60.072, -40.708) | -10.200 | 1.71E-18 | 1.98E-16 |
| negative regulation of leukocyte apoptotic process | -63.415 | 6.381 | (-75.922, -50.908) | -9.938 | 7.86E-18 | 7.28E-16 |
| sulfur compound metabolic process | -25.949 | 2.638 | (-31.119, -20.779) | -9.836 | 1.42E-17 | 1.16E-15 |
| proteoglycan metabolic process | -36.575 | 3.801 | (-44.025, -29.125) | -9.623 | 4.88E-17 | 3.77E-15 |
| leukocyte apoptotic process | -46.150 | 4.912 | (-55.778, -36.522) | -9.395 | 1.82E-16 | 1.13E-14 |
| neuron projection extension involved in neuron projection guidance | -42.305 | 4.533 | (-51.190, -33.420) | -9.333 | 2.61E-16 | 1.55E-14 |
| regulation of leukocyte apoptotic process | -48.549 | 5.204 | (-58.749, -38.349) | -9.328 | 2.67E-16 | 1.55E-14 |
| response to chemokine | -61.512 | 6.609 | (-74.466, -48.558) | -9.307 | 3.03E-16 | 1.66E-14 |
| glycoprotein metabolic process | -28.814 | 3.155 | (-34.998, -22.630) | -9.131 | 8.29E-16 | 3.94E-14 |
| negative regulation of axon extension involved in axon guidance | -43.733 | 4.843 | (-53.225, -34.241) | -9.030 | 1.48E-15 | 6.39E-14 |

\* Abbreviations: Std.Err: standard error; Adj.P: adjusted p-value. CI: confidence interval.
\* See Supplemental Digital Content Table S8 for multivariable model adjusted for CN-105, age, sex, education years, *ApoE*-e2 copy number, *ApoE*-e4 copy number, BMI, surgery type, ASA physical status classification, and baseline enrichment score.

### Unbiased Analysis of APOE4 carrier status and CN-105’s effects on Changes in Protein Intensity and Pathway Enrichment Score

After FDR adjustment there was no significant effect of *APOE4* carrier status, and no significant interaction effect of *APOE4* carrier status by time, on CSF protein levels (see Supplemental Digital Content Section 2 - *ApoE-e4* effect). Thus, *APOE4* carriers (as compared to non-carriers) did not have significant differences in baseline or 24 hour postoperative changes in CSF protein levels.

Next, we examined the effect of treatment with the APOE mimetic peptide CN-105 on postoperative CSF protein and pathway changes (see Supplemental Digital Content Table S3 for baseline characteristics of MARBLE participants who received CN-105 vs placebo). There was no significant effect of CN-105 on pre-operative to 24-hour post-operative CSF protein intensity level changes in univariable models (Supplemental Digital Content Fig S2, Table S4) or in multivariable models adjusted for numerous baseline patient characteristics and surgery type (Supplemental Digital Content Table S9, Supplemental Digital Content Data Sheet 8). In univariable models, the TM130 protein showed the largest magnitude upregulation by CN-105 (log_2_ FC = 0.74, SE = 0.48, 95% CI = [-0.20, 1.69], Supplemental Digital Content Data Sheet 10); and the SAA1 protein showed the largest magnitude down-regulation by CN-105 (log_2_ FC = - 0.78, SE = 0.27, 95% CI = [-1.30, -0.25]), though neither of these changes were significant after FDR correction. Further, there was no significant effect of CN-105 dose level (0, 0.1, 0.5 or 1 mg/kg) on postoperative protein intensity level changes in univariable models (see Supplemental Digital Content Table S5). There was also no significant effect of CN-105 on postoperative CSF protein pathway enrichment score changes in univariable models (see Supplemental Digital Content Fig. S3; Table S6) or in multivariable models adjusted for numerous baseline patient characteristics and surgery type (Supplemental Digital Content Table S10, Supplemental Digital Content Data Sheet 9).

### Results from the Replication Cohort

To corroborate these findings, we examined pre to 24-hour postoperative changes in CSF protein levels in an independent replication cohort of 96 participants from two other studies (see methods for replication cohort description; Supplemental Digital Content Section 3 for additional replication cohort information). Replication cohort baseline characteristics are provided in Supplemental Digital Content Table S11. Of the 418 proteins that were significantly upregulated after surgery in the MARBLE cohort (after FDR correction) and detectable in the replication cohort, ∼78% (327) were also significantly upregulated postoperatively (p-value<0.05) in the replication cohort. Further, of the 402 proteins that were significantly downregulated after surgery in the MARBLE cohort (after FDR correction) and detectable in the replication cohort, ∼64% (259) were also significantly downregulated postoperatively (p-value<0.05) in the independent replication cohort.

For the pathway effects, ∼58% (261 of the 447 pathways that were FDR-significantly upregulated in the MARBLE cohort) were also significantly upregulated in the replication cohort; and ∼62% (302 of the 485 pathways that were FDR-significantly down-regulated in the MARBLE cohort) were also significantly downregulated in the replication cohort. Further, there was a high degree of correlation between the effect sizes for, and directionality of, the postoperative changes in both the proteins and the pathways seen in the MARBLE cohort and the replication cohort (Figure 2A, Spearman’s rho =0.861 (95% CI: [0.823, 0.892], p<0.001; Figure 2B, Spearman’s rho =0.720 (95% CI: [0.680, 0.757], p-value<0.001, respectively).

**Figure 2.**
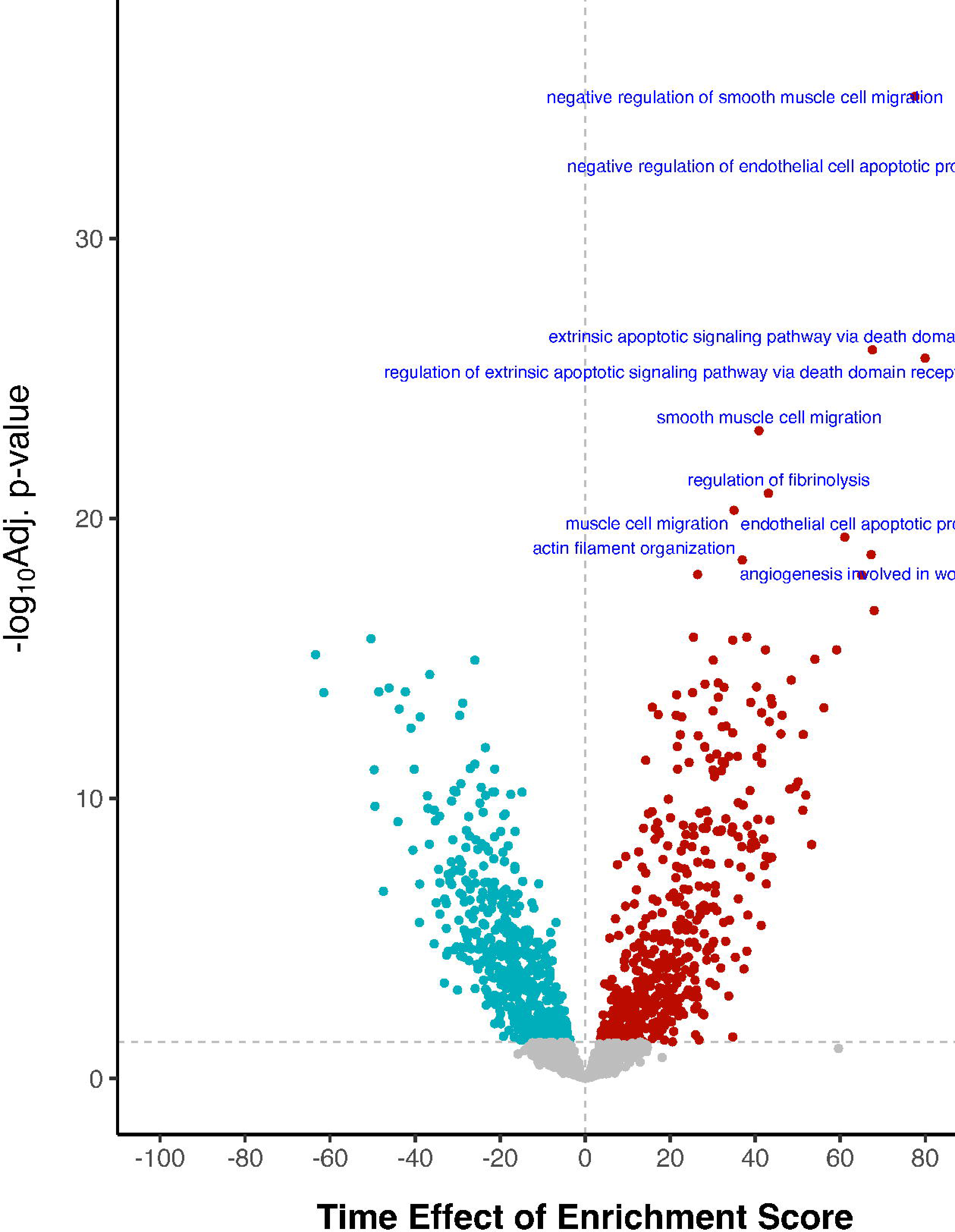
Spearman’s correlations (with 95% confidence interval) between postoperative changes in proteins and pathways between the MARBLE and replication cohorts. A) Scatterplot of the log2 FC of the 820 proteins (A) that showed a significant temporal effect in the MARBLE cohort (418 up-regulated proteins and 402 down-regulated proteins). The diagonal dashed line is the identity line. B) Scatterplot of enrichment score changes for 932 CSF pathways that showed an FDR-significant change from before to 24 hours after surgery in the MARBLE cohort (447 up-regulated pathways and 485 down-regulated pathways). The diagonal dashed line is the identity line. In both panels, red dots indicate proteins or pathways that were up- or down-regulated in both cohorts; green dots indicate proteins or pathways that were up- or down-regulated in the MARBLE cohort only. ES: Enrichment Score.

## DISCUSSION

This work demonstrates widespread and significant CSF proteome changes 24 hr after non-cardiac/non-neurologic surgery in a moderate sized cohort of older adults. Further, these postoperative CSF protein and pathway changes remained highly consistent across both univariable and multivariable adjusted analyses (to rule out confounding by patient characteristics or surgery type), and in CSF samples from an independent replication cohort. We identified postoperative changes in numerous CSF proteins and pathways involved in neuroinflammation and immune system activation, particularly in monocyte and neutrophil stress response pathways. We also observed postoperative changes in endothelial, actin and collagen pathways that could allow leukocytes to enter the CNS, and changes consistent with increased postoperative apoptosis of leukocytes, endothelial cells and/or other cell types within the CNS.

Surgical trauma is thought to lead to neuroinflammation via blood brain barrier dysfunction, microglial activation, and neuronal damage.^22^ Our results demonstrate substantial changes in CSF proteins within these pathways and processes at 24 hours after surgery. One of the most upregulated CSF proteins after surgery was serum amyloid A1 (SAA1), an acute phase response protein expressed in response to inflammation and tissue injury,^23^ such as following surgical stress. SAA1 can cross the blood-brain barrier and cause an increase in the number of microglia and pro-inflammatory factors in the brain,^24, 25^ and SAA1 protein levels also significantly increased in the hippocampus after surgery in mice.^25^ CSF plasminogen activator inhibitor-1 (PAI-1) levels were also significantly up-regulated following surgery here, consistent with the established role of glucocorticoid stress hormones in shifting the hippocampal balance between expression of Pal-1 and its target, tissue plasminogen activator (tPA).^26^ A cellular stress response is also consistent with the postoperative down-regulation of Collagen Type I Alpha 1 Chain (COL1A1) and Collagen Type I Alpha 2 Chain (COL1A2) observed here, since collagen expression by fibroblasts is downregulated by stress-induced adrenal cortisol release in a dose-dependent manner^27^ that can be blocked by glucocorticoid receptor (GR) antagonists.^28^ Overall, these data suggest that similar stress-induced protein level changes as those seen in these in non-CNS biological contexts also within the human central nervous system (ie CSF) of older adults following non-cardiac/non-neurologic surgery.

Consistent with the individual postoperative CSF collagen protein level decreases discussed above, the collagen fibril organization pathway was also significantly downregulated after surgery, which may facilitate the entry of peripheral leukocytes and monocytes into the CNS, as seen in both mouse surgical models^29–31^ and in older surgical patients.^32^ These mouse studies and the postoperative changes in CSF biological pathways described here are consistent with the idea that after surgical stress in older adults, leukocytes migrate from the vascular endothelium, cross blood vessel walls and the surrounding smooth muscle via diapedesis to reach the brain parenchyma and interstitial fluid, which is in equilibrium with the CSF. This process involves actin structure changes within the vascular endothelium, consistent with the postoperative changes in CSF actin filament organization pathways seen here.^33, 34^ After entering the central nervous system, these leukocytes (especially neutrophils and monocytes) likely undergo apoptosis due to their known short half-life (6-8 hours for neutrophils^35^), consistent with the postoperative CSF alterations of numerous apoptosis-related pathways observed here.

Indeed, of the top 40 CSF proteomic pathways that we found were upregulated after surgery, 8 (including 4 of the top 5) involved inflammation, immune response, and/or leukocyte/monocyte/neutrophil biology, 4 involved apoptosis, 7 involved wound healing/platelets/coagulation, and 6 involved actin/cytoskeleton biology. Six pathways downregulated after surgery were related to neuronal/synaptic function and development, and 19 were related to general growth/development (including the TGFβ pathway). Taken together, these data fit with a model in which the postoperative activation of inflammatory and stress pathways leads to decreased tissue development/growth processes (perhaps as a consequence of decreased TGF beta signaling) and decreased synaptic and neuronal function.

This study has several limitations. First, while we detected over 2,000 different CSF proteins, this also increased the burden of false discovery rate-based multiple testing correction, which may have reduced power to detect effects of either CN-105 or *APOE4* on CSF protein or pathway levels. Second, our moderate sample size (137 patients) relative to the number of proteins measured (>2,000) may also explain why we did not detect statistically significant effects (after FDR correction) of CN-105 treatment or *APOE4* carrier status. Thus, the lack of a detectable statistically significant effect of APOE4 on CSF protein levels, or of CN-105 on postoperative protein or pathway changes, should not be viewed as definitive evidence that neither of these effects exists. Third, like all unbiased mass spectrometry-based proteomic studies, the protein intensities studied here are semi-quantitative, not truly quantitative, measurements. Fourth, here we studied CSF proteomic changes at just one postoperative time point (i.e. 24 hours after surgery). A more detailed time course of postoperative CSF protein and pathway changes would provide greater insights into temporal change trajectories, though this would be challenging: it would require multiple postoperative lumbar punctures or intrathecal catheter placements.

This study also has several key strengths. These include: 1) a moderately large cohort of older surgical patients that underwent pre- and 24 hr post-operative lumbar punctures for research, 2) an independent replication cohort of CSF samples from older non-cardiac/non-intracranial surgery patients in other studies that also underwent pre- and 24 hr post-operative lumbar punctures; 3) comprehensive unbiased proteomic analysis and rigorous statistical analyses with FDR-based multiple comparison correction; and 4) pathway analysis using ssGSEA and multivariable models to control for potential confounders and covariates. Finally, a fifth strength of this work is that even though there were significant technical differences in the mass spectroscopy assays used for the MARBLE cohort samples versus the independent replication cohort samples, we nonetheless observed very similar postoperative changes in CSF protein and pathway levels in both cohorts, providing strong confidence in the overall validity of these results.

This work provides valuable insights into the CSF proteomic and pathway changes that occur in response to surgery in older adults, and represents one of the first systematic and unbiased descriptions of the postoperative molecular changes that take place within the central nervous system of older adults who underwent non-cardiac/non-neurologic surgery. Further, the postoperative protein pathway changes seen here are consistent with prior animal model work and suggest that there is a postoperative leukocyte influx across the neurovascular endothelium into the brain parenchyma, the brain’s interstitial fluid and the CSF. Future studies should evaluate the role of these CSF protein and pathway changes in perioperative neurocognitive disorders among older adults, and test whether blocking specific postoperative protein or pathway changes within the human central nervous system helps prevent these disorders.

## List of abbreviations

CSF -: cerebrospinal fluid;
CNS -: central nervous system;
APOE -: apolipoprotein E;
MARBLE -: Modulating APOE to Reduce Brain infLammation and postoperative dElirium;
FDR -: false discovery rate;
ssGSEA -: single-sample gene set enrichment analysis;
GO-BP -: Gene Ontology database for biological processes.

## Declarations

### Ethics approval and consent to participate

Because Duke University owns intellectual property rights to CN-105, this study was reviewed by the Duke Office of Conflict Management, which determined that the study could be conducted at Duke under the auspices of an external IRB. Thus, MARBLE was approved by the Western Institutional Review Board and registered with clinicaltrials.gov prior to study initiation (ct.gov identifier: NCT03802396). MADCO-PC and INTUIT were approved by the Duke IRB (neither of these studies utilized CN-105), and registered with clinicaltrials.gov (NCT01993836, NCT03273335). All patients in each of these studies completed written informed consent prior to participation in any study activities.

### Consent for publication

Not applicable.

### Availability of data and materials

Raw data and sample-linking metadata from the MARBLE study, as well as protein .fasta file, Spectronaut spectral library and .SNE files, and protein group abundance table, have been uploaded to the ProteomeXchange consortium (PXD068209) via the MassIVE repository (ftp://MSV000099094@massive-ftp.ucsd.edu) and can be accessed using reviewer password MARBLECSF. Raw data from the replication cohort will be made available upon separate publication of that dataset or upon reasonable request.

### Authors’ contributions

Study concept and design: MWL, MWF, MCW, DL, MJD, MB

Proteomic Data Collection: MWF, MJV.

Statistical analysis: HZ, MWL, MWF, SL.

Data interpretation: HZ, MWL, MWF, SL, DL, MJD, MB.

Manuscript preparation: HZ, MWL, MWF, MCW, MJD, MB

Manuscript review and final approval: all authors

## Supporting information

Supplemental Digital Content - Word

Supplemental Digital Content - Excel

## Data Availability

Raw data and sample-linking metadata from the MARBLE study, as well as protein .fasta file, Spectronaut spectral library and .SNE files, and protein group abundance table, have been uploaded to the ProteomeXchange consortium (PXD068209) via the MassIVE repository (ftp://MSV000099094@massive-ftp.ucsd.edu) and can be accessed using reviewer password MARBLECSF.

ftp://MSV000099094@massive-ftp.ucsd.edu

## Acknowledgements

We thank Drs Beth Stevens, Nader Morshed and Matthew Johnson for helpful comments on this manuscript, and the older surgical patients who participated in the studies that provided the samples for this work.

## Collaborators

The MADCO-PC Study Group Also Includes: Brian E. Brigman, Jeffrey N. Browndyke, W. Michael Bullock, Jessica Carter, Joseph Chapman, Brian Colin, Mary Cooter Wright, Thomas A. D’Amico, James K. DeOrio, Ramon M. Esclamado, Michael N. Ferrandino, Jeffrey Gadsden, Grant E. Garrigues, Stuart Grant, Jason Guercio, Dhanesh Gupta, Ashraf Habib, David H. Harpole, Mathew G. Hartwig, Ehimemen Iboaya, Brant A. Inman, Anver Khan, Sandhya Lagoo-Deenadayalan, Paula S. Lee, Walter T. Lee, John Lemm, Howard Levinson, Christopher Mantyh, Joseph P. Mathew, David L. McDonagh, John Migaly, Suhail K. Mithani, Eugene Moretti, Judd W. Moul, Mark F. Newman, Brian Ohlendorf, Alexander Perez, Andrew C. Peterson, Glenn M. Preminger, Quintin Quinones, Cary N. Robertson, Sanziana A. Roman, Scott Runyon, Aaron Sandler, Faris M. Sbahi, Randall P. Scheri, S. Kendall Smith, Leonard Talbot, Julie K. M. Thacker, Jake Thomas, Betty C. Tong, Steven N. Vaslef, Nathan Waldron, Xueyuan Wang, and Christopher Young.

The INTUIT Study Group Also Includes: Leah Acker, Cindy Louise Amundsen, Oke Anakwenze, Harel Anolick, David Attarian, Pallavi Avasarala, Chakib Ayoub, Matthew Barber, Rachel Beach, Andrew Berchuck, Dan G. Blazer III, Michael Bolognesi, Rachele Brassard, Brian Brigman, William Michael Bullock, Thomas Bunning, Victor Cai, Yee Ching Vanessa Cheong, Soren K Christensen, Brian Colin, Mary Cooter Wright, Mitchell Wayne Cox, Thomas D’Amico, Brittany Anne Davidson, James Keith Deorio, Mark E. Easley, Sarada Eleswarpu, Detlev Erdmann, Mariana Feingold, Michael Nicolo Ferrandino, Jeffrey Gadsden, Mark Gage, Arun Ganesh, Grant Edward Garrigues, Rachel Adams Greenup, Ashraf Habib, Ashley Hall, Rhett K. Hallows, David Harpole Jr, Matthew Hartwig, Laura Havrilesky, Courtney Holland, Scott Thomas Hollenbeck, Thomas Hopkins, Edward Ross Houser ll, Samuel Huang, Ehimemen Iboaya, Brant Inman, William Jiranek, Russel Kahmke, Amie Kawasaki, Brendan Kelleher, Jay Han Kim, Jacob Klapper, Christopher Klifto, Rebecca Klinger, Stuart Knechtle, Sandhya A. Lagoo-Deenadayalan, Billy Lan, Walter Lee, Howard Levinson, Brian Lewis, Michael Lipkin, Christopher Mantyh, Hector Martinez-Wilson, John Migaly, Eugene Moretti, Judd Moul, Michael Muehlbauer, David Murdoch, Thomas L. Novick, Kathryn Odom, Brian Ohlendorf, Steven Olson, Deborah Oyeyemi, Shannon Page, Theodore Pappas, John Park, Andrew Peterson, Andreea Podgoreanu, Thomas J Polascik, Dana Portenier, Glenn M. Preminger, Rebecca Ann Previs, Edward Nandlal Rampersaud Jr, Melody Reese, Kenneth Roberts, Cary N. Robertson, Sanziana Alina Roman, Jason Rothman, Aaron Sandler, Siddharth Sata, Charles Scales Jr, Randall Scheri, Thorsten Seyler, Keri Anne Seymour, Nazema Y. Siddiqui, Shayan Smani, Michael Stang, Samuel David Stanley, Katherine Sweeney, Ayesha Syed, Martin V. Taormina, Julie Thacker, Jake Thomas, Betty Tong, Yanne Toulgoat-Dubois, Keith Vandusen, Nathan Waldron, Alison Weidner, Kent Weinhold, Samuel Wellman, David Williams, Marty Woldorff, Rosa Yang, Christopher Young, Sabino Zani, and Mimi Zhang.

The MARBLE Study Group Also Includes: Samuel B Adams, Cindy L Amundsen, Pallavi Avasarala, Chakib M. Ayoub, Matthew D Barber, Andrew Berchuck, Daniel G Blazer III, Kaj Blennow, Michael P. Bolognesi, Piper C. Boykin, Rachele Brassard, Brian E Brigman, Jeffrey N. Browndyke, Thomas Bunning, Victor Cai, Soren K Christensen, Mary Cooter Wright, Mitchel W Cox, Brittany A Davidson, James K DeOrio, Sarada Eleswarpu, Jacqueline M. Emerson, Detlev Erdmann, Melissa M Erickson, Bonita L Funk, Jeffrey Gadsden, Mark J Gage, Jeff R Gingrich, Rachel A Greenup, Christine Ha, Ashraf Habib, Ralph Abi Hachem, Ashley E Hall, Matthew G Hartwig, Laura J Havrilesky, Mitchell T Heflin, ; J. Taylor Herbert, Courtney Holland, Scott T Hollenbeck, Thomas J Hopkins, Bethany J. Hsia, Janet L. Huebner, Brant A Inman, David W Jang, William A. Jiranek, Russel R Kahmke, Isaac Karikari, Amie Kawasaki, Jacob A Klapper, Christopher S Klifto, Rebecca Klinger, Stuart J Knechtle, Sandhya A Lagoo-Deenadayalan, Walter T Lee, Howard Levinson, Brian D Lewis, Zhong Li, Michael E Lidsky, Michael E Lipkin, Christopher R Mantyh, Shelley R McDonald, John Migaly, Timothy E Miller, Suhail K Mithani, Eugene W. Moretti, Paul J Mosca, Judd W Moul, Thomas L Novick, Steven A Olson, Theodore N Pappas, John J Park, Andrew C Peterson, Brett T Phillips, Thomas J Polascik, Peter Potash, Glenn M Preminger, Rebecca A Previs, Melody Reese Smith, Kenneth Roberts, Cary N Robertson, Frank Rockhold, David Ryu, Charles D Scales Jr, Kevin N Shah, Randall P Scheri, Nazema Y Siddiqui, Shayan Smani, Kevin W Southerland, Michael T Stang, Ayesha Syed, Alicja Szydlowska, Julie K M Thacker, Niccolò Terrando, Noah J. Timko, Yanne Toulgoat-DuBois, Keith W. VanDusen, Anthony G Visco, Alison C Weidner, Marty Woldorff, Mamata Yanamadala, Sabino Zani Jr., and Henrik Zetterberg.

## Supplementary Material

Supplemental Digital Content – Word.docx

Supplemental Digital Content – Excel.xlsx

Dataset S1.xlsx; Dataset S2.xlsx; Dataset S3.xlsx

---

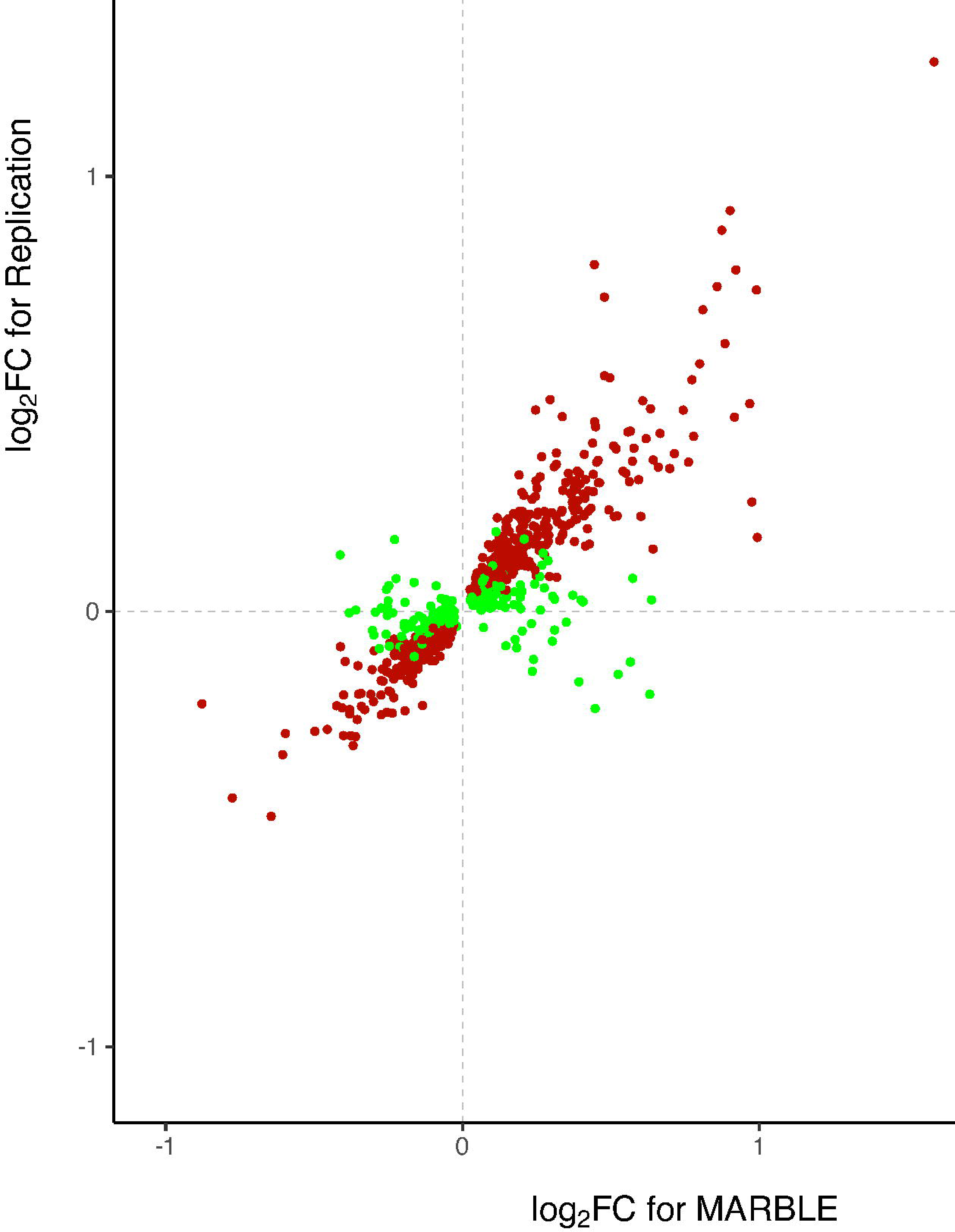

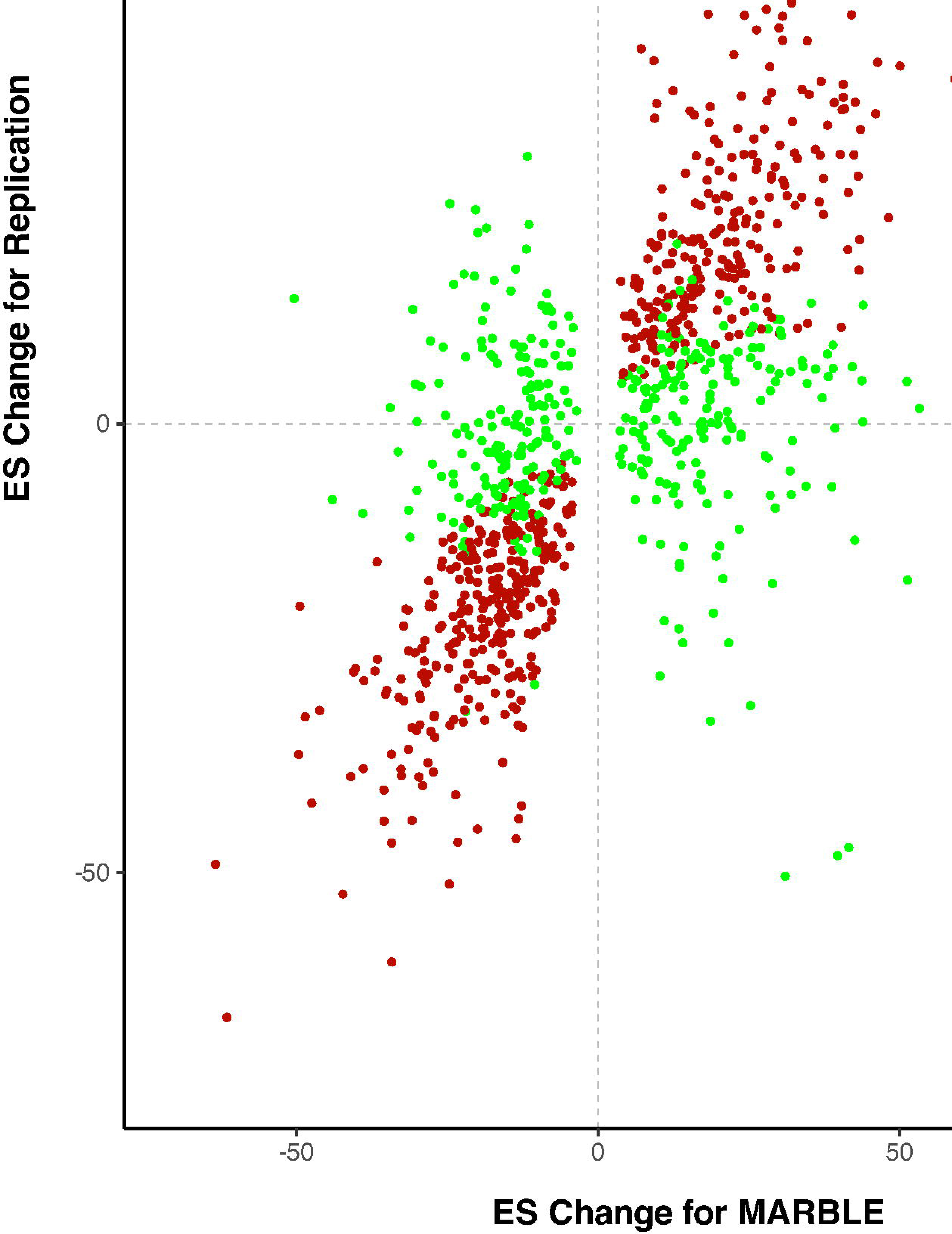

## REFERENCES

1. Evered L, Silbert B, Knopman DS, et al. Recommendations for the nomenclature of cognitive change associated with anaesthesia and surgery—2018. Anesthesiology 2018; 129(5):872–879.

2. Berger M, Terrando N, Smith SK, et al. Neurocognitive function after cardiac surgery: from phenotypes to mechanisms. Anesthesiology 2018; 129(4):829–851.

3. Vlisides P, Mashour GA. Perioperative stroke. Can J Anaesth 2016; 63(2):193.

4. Nobuhara CK, Bullock WM, Bunning T, et al. A protocol to reduce self-reported pain scores and adverse events following lumbar punctures in older adults. J Neurol 2020; 267:2002–2006.

5. VanDusen KW, Li Y-J, Cai V, et al. Cerebrospinal fluid proteome changes in older non-cardiac surgical patients with postoperative cognitive dysfunction. J Alzheimers Dis 2021; 80(3):1281–1297.

6. Higginbotham LA, Ping L, Dammer EB, et al. Proteomics identifies CSF biomarker panels reflective of pathological networks in the Alzheimer’s disease brain: Biomarkers (non neuroimaging): Novel biomarkers. Alzheimers Dement 2020; 16:e042227.

7. Johnson EC, Dammer EB, Duong DM, et al. Large-scale proteomic analysis of Alzheimer’s disease brain and cerebrospinal fluid reveals early changes in energy metabolism associated with microglia and astrocyte activation. Nat Med 2020; 26(5):769–780.

8. Bartels K, Li Y-J, Li Y-W, et al. Apolipoprotein epsilon 4 genotype is associated with less improvement in cognitive function five years after cardiac surgery: a retrospective cohort study. Canadian journal of anaesthesia= Journal canadien d’anesthesie 2015; 62(6):618.

9. Schenning KJ, Murchison CF, Mattek NC, et al. Surgery is associated with ventricular enlargement as well as cognitive and functional decline. Alzheimer’s & Dementia 2016; 12(5):590–597.

10. Berger M, Cooter M, Roesler AS, et al. APOE4 Copy Number-Dependent Proteomic Changes in the Cerebrospinal Fluid 1. J Alzheimers Dis 2021; 79(2):511–530.

11. Ayton S, Janelidze S, Roberts B, et al. Acute phase markers in CSF reveal inflammatory changes in Alzheimer’s disease that intersect with pathology, APOE ε4, sex and age. Prog Neurobiol 2021; 198:101904.

12. Lei B, James ML, Liu J, et al. Neuroprotective pentapeptide CN-105 improves functional and histological outcomes in a murine model of intracerebral hemorrhage. Sci Rep 2016; 6(1):34834.

13. Laskowitz DT, Van Wyck DW. ApoE mimetic peptides as therapy for traumatic brain injury. Neurotherapeutics 2023; 20(6):1496–1507.

14. Guptill JT, Raja SM, Boakye Agyeman F, et al. Phase 1 randomized, double blind, placebo controlled study to determine the safety, tolerability, and pharmacokinetics of a single escalating dose and repeated doses of CN 105 in healthy adult subjects. The Journal of Clinical Pharmacology 2017; 57(6):770–776.

15. Timko NJ, Cooter Wright M, Reese M, et al. The ApoE Mimetic Peptide CN-105 in Older Surgical Patients: The MARBLE Phase 2 Randomized Clinical Trial. JAMA Surgery 2025; Under Review.

16. Berger M, Oyeyemi D, Olurinde MO, et al. The INTUIT study: investigating neuroinflammation underlying postoperative cognitive dysfunction. J Am Geriatr Soc 2019; 67(4):794–798.

17. Berger M, Browndyke JN, Cooter Wright M, et al. Postoperative changes in cognition and cerebrospinal fluid neurodegenerative disease biomarkers. Ann Clin Transl Neurol 2022; 9(2):155–170.

18. Benjamini Y, Yekutieli D. The control of the false discovery rate in multiple testing under dependency. Annals of statistics 2001:1165–1188.

19. Subramanian A, Tamayo P, Mootha VK, et al. Gene set enrichment analysis: a knowledge-based approach for interpreting genome-wide expression profiles. Proc Natl Acad Sci 2005; 102(43):15545–15550.

20. Barbie DA, Tamayo P, Boehm JS, et al. Systematic RNA interference reveals that oncogenic KRAS-driven cancers require TBK1. Nature 2009; 462(7269):108–112.

21. Schork K, Podwojski K, Turewicz M, et al. Important issues in planning a proteomics experiment: statistical considerations of quantitative proteomic data. Quantitative Methods in Proteomics: Springer; 2021:pp. 1–20.

22. Alam A, Hana Z, Jin Z, et al. Surgery, neuroinflammation and cognitive impairment. EBioMedicine 2018; 37:547–556.

23. Sun L, Ye RD. Serum amyloid A1: Structure, function and gene polymorphism. Gene 2016; 583(1):48–57.

24. Gao Y, Tang S, Liu J, et al. The Administration of Circulating Extracellular Vesicles Modified by Anesthesia and Surgery Induces Delirium-Like Behaviors in Aged Mice. CNS Neurosci Ther 2025; 31(6):e70483.

25. Jang WY, Lee BR, Jeong J, et al. Overexpression of serum amyloid a 1 induces depressive-like behavior in mice. Brain Res 2017; 1654(Pt A):55–65.

26. Mennesson M, Revest JM. Glucocorticoid-Responsive Tissue Plasminogen Activator (tPA) and Its Inhibitor Plasminogen Activator Inhibitor-1 (PAI-1): Relevance in Stress-Related Psychiatric Disorders. Int J Mol Sci 2023; 24(5).

27. Chae M, Bae IH, Lim SH, et al. AP Collagen Peptides Prevent Cortisol-Induced Decrease of Collagen Type I in Human Dermal Fibroblasts. Int J Mol Sci 2021; 22(9).

28. Mi Y, Wang W, Zhang C, et al. Autophagic Degradation of Collagen 1A1 by Cortisol in Human Amnion Fibroblasts. Endocrinology 2017; 158(4):1005–1014.

29. Yang T, Xu G, Newton P, et al. Maresin 1 attenuates neuroinflammation in a mouse model of perioperative neurocognitive disorders. Br J Anaesth 2019; 122(3):350–360.

30. Terrando N, Eriksson LI, Kyu Ryu J, et al. Resolving postoperative neuroinflammation and cognitive decline. Ann Neurol 2011; 70(6):986–995.

31. Hirsch J, Vacas S, Terrando N, et al. Perioperative cerebrospinal fluid and plasma inflammatory markers after orthopedic surgery. J Neuroinflammation 2016; 13:1–12.

32. Berger M, Murdoch DM, Staats JS, et al. Flow cytometry characterization of cerebrospinal fluid monocytes in patients with postoperative cognitive dysfunction: a pilot study. Anesthesia & Analgesia 2019; 129(5):e150–e154.

33. Heemskerk N, Schimmel L, Oort C, et al. F-actin-rich contractile endothelial pores prevent vascular leakage during leukocyte diapedesis through local RhoA signalling. Nat Commun 2016; 7(1):10493.

34. Wójciak-Stothard B, Williams L, Ridley AJ. Monocyte adhesion and spreading on human endothelial cells is dependent on Rho-regulated receptor clustering. J Cell Biol 1999; 145(6):1293.

35. Summers C, Rankin SM, Condliffe AM, et al. Neutrophil kinetics in health and disease. Trends Immunol 2010; 31(8):318–24.

