## Supplemental Digital Content - Word for "Effects of Non-Cardiac/Non-neurologic Surgery and Anesthesia on the CSF Proteome, and Modulation by the APOE Mimetic Peptide CN-105"

**Section 1: Methods for Processing and Acquisition of the MARBLE and Replication Cohort CSF Proteomic Data**

***MARBLE Trial Description:***

Modulating APOE to Reduce Brain infLammation and postoperative dElirium (MARBLE), was a single center, escalating dose, FDA-regulated, triple blind phase II study designed to test the safety of the investigational product CN-105 for the prevention of postoperative delirium among older (ie age ≥60 years old) non-cardiac, non-intracranial surgery patients. MARBLE was triple blinded- the patients and their family members, clinical teams caring for the patients, and the research team were all blinded to randomization assignment. Randomization was performed according to an electronic randomization list designed before the start of MARBLE study enrollment, via a permuted block design (N=8 block size plus a single randomly placed N=3 block).

MARBLE study patient enrollment occurred between April 17, 2019, and December 28, 2022. Eligible individuals were English-speaking patients aged ≥60 years who were scheduled for a ≥2-hour major non-cardiac/non-intracranial surgery with a planned overnight hospital stay. Patients were excluded from the study if they were incarcerated, if they planned to undergo systemic chemotherapy between the baseline/preoperative and 6-week postoperative study visits, if they were unable to undergo lumbar punctures (i.e. due to anticoagulant drugs), if they experienced head trauma between the time of their baseline visit and their 6 week postoperative study visit, or if they were inappropriate for study inclusion per the judgement of the study PI.^16^ There were no preoperative cognitive function thresholds or exclusions used to define which patients could enroll in MARBLE.

MARBLE enrolled three successive cohorts of 67 patients- in the first cohort, 50 patients were randomized to receive CN-105 at 0.1 mg/kg and 17 were randomized to receive placebo, starting just before the start of surgery and then continuing every 6 hours after surgery for up to 3 days (13 doses) or hospital discharge, whichever occurred first. In the second cohort, 50 patients were randomized to receive CN-105 at 0.5 mg/kg and 17 were randomized to receive placebo; in the third cohort, 50 patients were randomized to receive CN-105 at 1.0 mg/kg and 17 were randomized to receive placebo- the drug (or placebo) was administered on the same schedule in cohorts two and three as in cohort one. Thus MARBLE was designed so that at study completion, there would be ~50 patients who received placebo, ~50 who received 0.1 mg/kg of CN-105, ~50 who received 0.5 mg/kg of CN-105, and ~50 who received 1.0 mg/kg of CN-105.

Patients’ clinical teams determined their hospital discharge date/time without regard to study participation; thus, patients were not kept in the hospital longer than clinically necessary for research purposes. The Duke Investigational Pharmacy prepared CN-105 or placebo in plastic IV bags covered with opaque shrouds, which appeared identical whether they contained CN-105 or placebo, and the drug or placebo was given to patients by the clinical teams caring for the study patients, per the per the dosing schedule discussed above.

The pre-specified primary outcome of the MARBLE trial was safety, defined as the rate of ≥ grade 2 adverse events (AEs) that occurred per patients, in terms of both the incidence of ≥ grade 2 AEs and number of these AEs per patient among CN-105- vs placebo-randomized patients after surgery through the 6-week study follow-up period.^15^ AEs were defined according to version 5 of the Common Terminology Criteria for Adverse Events (CTCAEv5). AEs were tracked from participant reports and via review of participants’ clinical records by study physicians who were blinded to randomization group, from the time of study drug administration until the time of the 6-week postoperative study visit.

Pre-specified secondary outcomes of the MARBLE trial included the pre to 24 hr postoperative change in the cytokines IL-6, IL-8, G-CSF and MCP-1, delirium incidence and severity, and cognitive function at 6 weeks after surgery. The primary MARBLE study outcome and these secondary study outcomes of the MARBLE trial have been published separately.^15”^

This manuscript uses samples from the MARBLE trial (and two other cohort studies- MADCO-PC and INTUIT) to study the effects of surgery and anesthesia on changes in the CSF proteome from before to 24 hours after non-cardiac/non-intracranial surgery among older adults. Thus, this manuscript is effectively a discovery-based study that uses samples from a randomized controlled trial and two cohort studies; the proteins levels studied here were not a pre-specified outcome measure in any of these individual studies.

Nonetheless, since brain-related complications (ie delirium, cognitive dysfunction, stroke, etc) are (as a class) the most common postsurgical complications among older adults, a fundamental questions is: what are the molecular effects of surgery on other organ systems (outside the central nervous system or CNS) on the aging human brain and central nervous system? The data presented here answer this question in rigorous fashion using unbiased mass spectroscopy-based proteomics on pre and postoperative CSF samples, with strict multiple comparison correction, and an independent replication cohort.

**Processing of MARBLE study samples.** Cryovials were arranged in the order given in Supplemental Data **Sheet S2**, thawed at 25 °C and kept on ice, and 0.5 mL of each sample was transferred to 0.75 mL Matrix tubes (ThermoFisher Scientific). Aliquots of a study pool QC (SPQC) were created by mixing equal volumes (50 µL) of samples from the first plate, and 0.5 mL aliquots were transferred to Matrix tubes. Tubes were arranged in a 96-well format**,** capped lightly with a TPE Lyo Cap-96 (Micronic) and frozen overnight at -80 °C. Next, protein concentrations were measured using 5 μL of each sample by Bradford assay (**see Dataset S1**). The next day, samples were lyophilized overnight to dryness and stored at -80 °C.

For trypsin digestion, all buffers were prepared fresh. To each lyophilized CSF sample, we added 120 μL of 8M Urea and 10 mM DTT in 100 mM ammonium bicarbonate (AmBic). Samples were then incubated for 45 mins at 32 °C at 100 rpm on a Thermomixer R (Eppendorf) set at 1000 rpm. Next, 10 μL of 0.25 M iodoacetamide (IAM) in 50 mM AmBic was added to each sample, followed by incubation at room temperature for 30 min in the dark. Next, 330 µL AmBic and 10 mM DTT in AmBic were added, and then 50 μL of 0.4 µg/μL SequENZ® Trypsin (Worthington) in AmBic was added. All samples were allowed to incubate with shaking at 32 °C on a Thermomixer for 2 h. Finally, the digestion was quenched with addition of 50 μL 5% (v/v) trifluoroacetic acid (TFA) in water.

Samples were desalted using OASIS HLB 96-well plates (30 μm particle size 30 mg bed; Waters Corporation) using vacuum filtration. Plates were pre-eluted with 0.5 mL of 49.9/50/0.1 (v/v/v) water/acetonitrile/formic acid (H_2_O/MeCN/FA) and equilibrated with 2 x 0.5 mL of 0.1% FA. Samples were added to the plate slowly under vacuum and washed 2x with 0.5 mL of 0.1% FA. Samples were eluted from the SPE Bed into 0.75 mL Matrix tubes using 300 μL of 49.9/50/0.1 (v/v/v) H_2_O/MeCN/FA, followed by 300 µL of 99.0/0.1 (v/v) MeCN/FA and dried under N_2_ for approximately 1 h at 30 °C (SPEDry 96). Once volume was reduced by greater than half, samples were frozen at -80 °C and lyophilized to dryness as previously described. Finally, samples were each resuspended in 80 µL of 1/2/97 (v/v/v) TFA/MeCN/H_2_O containing 40 fmol/µL MassPREP yeast alcohol dehydrogenase 1 (ADH1; Waters), centrifuged and transferred to Maximum Recovery LC Vials (Waters).

**Quantitative Data-Independent-Acquisition (DIA) LC-MS/MS Analysis of MARBLE study samples.** Tryptic digests were analyzed by microflow LC-MS/MS using an ACQUITY UPLC (Waters Corporation) interfaced to an Exploris 480 Orbitrap (ThermoFisher Scientific) using a Optamax NG source with heated electrospray probe. Briefly,10 μL of peptide digests were separated on a 1 mm x 10 mm 1.7 μm ACQUITY Premier CSH C18 column (Waters) using a flow rate of 100 μL/min, a column temperature of 55 °C and a gradient of 3-28% (v/v) MeCN:H2O containing 0.1% FA over 60 min with source parameters: sheath gas, 23; aux gas, 5; sweep gas, 0; spray voltage, 3.5 kV; capillary temperature, 300 °C; aux gas heater temp, 125 °C. An M-Class Auxiliary Solvent Manager (Waters) was utilized to perform post-column addition of 50/50 (v/v) MeCN/DMSO at a flow rate of 6 µL/min.^1^

The data independent analysis (DIA) used a staggered, overlapping window method^2^ with a 120,000 resolution precursor ion (MS1) scan from 390-1010 m/z, AGC target of 1000% and maximum injection time (IT) of 60 ms and RF lens of 40%; data was collected in centroid mode. MS/MS was performed using targeted MS2 (tMS2) method with default charge state = 3, 30,000 resolution, automatic gain control (AGC) target of 1000% and maximum ion injection time (max IT) of 60 ms, and a normalized collision energy (NCE) of 30; data was collected in centroid mode. The DIA windows were generated using EncyclopeDIA (<https://bitbucket.org/searleb/encyclopedia/wiki/Home>) with a mass range of 400-1000 and 45 x 28 m/z windows, with a 22 window cycle. The cycle time between MS1 scans was 1.87 s, and the total injection-to-injection time was 67 min.

Raw MS data was demultiplexed and converted to .htrms format using HTRMS converter (v. 17.2.230208.55965; Biognosys) and processed in Spectronaut 17 (17.2.230208.55965; Biognosys). Searches used a Uniprot *Homo sapiens* consensus database (downloaded on 10/16/2020) and appended with sequences of bovine trypsin, yeast ADH1, and a concatemer of single amino acid variants (20,391 total entries). Search settings were as previously described,^3^ including N-terminal semitrypsin/P specificity, up to 2 missed cleavages and fixed carbamidomethyl (Cys). For DIA analysis, default extraction, calibration, identification, and protein inference settings were used. Data was filtered at a 1% precursor and protein group false discovery rate (q-value). All precursors that met a q-value were used for local normalization based on Callister et al.^4^, and protein abundances were calculated using the MaxLFQ algorithm.^5^ Two duplicate samples from the MARBLE study were averaged together after log_2_ transformation.

**Processing of replication cohort samples.** Twenty microliters of 8M urea in 50 mM Tris, pH 8 was added to 96-well “digestion plates” (Eppendorf 96/500 μl Protein Lobind) and dried in a SpeedVac. Cryovials were arranged in a 96-well format in the pre-randomized order. Samples were thawed at 25 °C and kept on ice, and 20 μL of each sample was transferred to the digestion plate. A Quick Start Bradford Protein Assay (Bio-Rad) was performed on an additional 5 μL CSF. Aliquots of a study pool QC (SPQC) were created by mixing equal volumes (5 µL) of all samples. Plates were frozen overnight at -80 °C.

The digestion plates were sealed with foil, and protein was denatured by mixing on a Thermomixer using a deep-well plate (DWP500) adapter at 25 °C for 20 min at 500 rpm. Reduction and alkylation were performed on a Thermomixer with a DWP500 and heated lid by addition 20 μL of 15 mM Tris(2-carboxyethyl)phosphine (TCEP) and 60 mM chloroacetamide (CAA) in 50 mM AmBic at 32 °C and 500 rpm for 20 min. Next, 10 μL of Worthington Lys-C (0.05 µg/µL in AmBic) were added and incubated at 32 °C and 500 rpm for 2 h. Samples were further diluted with 30 μL of 50 mM AmBic and 20 μL of Sequenz trypsin (0.05 µg/µL in AmBic) were added. After 2 h at 32 °C, 11 μL of neat trifluoroacetic acid was added to a final concentration of 1% per volume. Finally, 10 μL of acid-quenched digests were loaded directly on EvoTips containing 40 μL of 0.1% formic acid (buffer A) and 100 fmol ADH using a ViaFlow-96 pipettor (Integra). EvoTips were further washed with 3 x 100 μL of buffer A. Finally, a post-digestion SPQC was created by mixing 10 μL of all digests from plate 1.

**Quantitative Data-Independent-Acquisition (DIA) LC-MS/MS Analysis of replication cohort samples.** Quantitative LC/MS-MS was performed using 10 µL of each sample and replicates of an SPQC pool (either pooled pre or post digestion), using an Evosep One LC interfaced to a ThermoFisher Orbitrap Astral. The Evosep used a 30 sample-per-day method with a Bruker Pepsep 15 cm x 150 μm column (1.5 μm particle size) and a PepSep Sprayer and stainless steel (30 μm) emitter. Data collection on the Orbitrap Astral mass spectrometer was performed in DIA mode of acquisition with a 240,000 resolution full MS scan every 0.6 s from m/z 380-980 with an AGC target of 500%. DIA MS/MS scans were acquired in the Astral analyzer using fixed windows of 4 m/z from m/z 380-80 m/z, target AGC of 500% and max fill time of 6 ms. A normalized collision energy of 28% was used for all MS2 scans.

Raw MS data was converted to *.htrms format using HTRMS converter and processed in Spectronaut 18 (18.7.240325.55695 Biognosys). A spectral library was built using direct-DIA searches which used a *Homo sapiens* consensus database downloaded on 4/22/24 using FragPipe and appended contaminant sequences, Lys-C, bovine trypsin, yeast ADH1, human ApoL1 C-terminal variants and a concatemer of common variants. Pulsar searches were performed on all individual samples, and identification and quantification were performed as described above.

**Supplemental Data.** Raw data and sample-linking metadata from the MARBLE study, as well as protein .fasta file, Spectronaut spectral library and .SNE files, and protein group abundance table, have been uploaded to the ProteomeXchange consortium (PXD068209) via the MassIVE repository (ftp://MSV000099094@massive-ftp.ucsd.edu) and can be accessed using reviewer password MARBLECSF. Raw data from the replication cohort will be made available upon separate publication of that dataset or upon reasonable request.

**Missing data Imputation.**

The missing data in the CSF proteome are due to zero precursors of the protein in the mass spectrometry assay. The missing protein intensity levels were imputed using a random forest in both the MARBLE cohort and replication cohort. The MARBLE dataset consists of 289 samples (including SPQC and replicate samples), in which 2,162 proteins were detected. We imputed missing intensities of the proteins using the random forest method, which has an outstanding performance as compared to other imputation techniques for high-dimensional data. We included 2,086 proteins with a missing proportion ≤ 80% at both the pre-operative and post-operative timepoints (i.e., we excluded 76 proteins that had a missing proportion > 80%). The missing data imputation using the random forest was achieved via averaging unpruned regression trees. We imputed the pre-operative and post-operative data simultaneously to account for the within-subject correlation in protein levels. We used 100 trees and 10 iterations and computed out-of-bag error (OOB) as a representation of the true imputation error. Imputation performance of protein intensity levels using random forest showed an out-of-bag error (OOB), measured by the normalized root mean squared error (NRMSE) of 0.189, suggesting satisfactory performance.^6^

The replication cohort had a total of 3223 proteins and 226 proteomics samples at the pre-operative and post-operative timepoint. We included 2871 proteins with a missing proportion ≤ 80% at both the pre-operative and post-operative timepoints. We removed MARBLE study participants in the replication cohort, resulting in a total of 192 proteomics samples (96 participants). We imputed the missing intensity levels using the same approach as in the MARBLE cohort. The imputation performance in the replication cohort, measured by NRMSE, was 0.159, suggesting a satisfactory performance.

**REFERENCES**

1.	Distler U, Lacki MK, Schumann S, et al. Enhancing Sensitivity of Microflow-Based Bottom-Up Proteomics through Postcolumn Solvent Addition. *Anal Chem* 2019; 91(12):7510-7515.

2.	Pino LK, Just SC, MacCoss MJ, et al. Acquiring and Analyzing Data Independent Acquisition Proteomics Experiments without Spectrum Libraries. *Mol Cell Proteomics* 2020; 19(7):1088-1103.

3.	Yurashevich M, Devinney M, Foster MW, et al. Cerebrospinal fluid proteome of patients with persistent pain and/or postpartum depression after elective cesarean delivery: An exploratory prospective cohort study. *J Clin Anesth* 2025; 104:111855.

4.	Callister SJ, Nicora CD, Zeng X, et al. Comparison of aerobic and photosynthetic Rhodobacter sphaeroides 2.4.1 proteomes. *J Microbiol Methods* 2006; 67(3):424-36.

5.	Cox J, Hein MY, Luber CA, et al. Accurate proteome-wide label-free quantification by delayed normalization and maximal peptide ratio extraction, termed MaxLFQ. *Mol Cell Proteomics* 2014; 13(9):2513-26.

6.	Stekhoven DJ, Bühlmann P. MissForest—non-parametric missing value imputation for mixed-type data. *Bioinformatics* 2012; 28(1):112-118.

**Section 2: Additional Methods and Results from the MARBLE Cohort**

**Table S1**. The effect of surgery type on protein intensity change from before to 24 hours after surgery, for the top 10 proteins that showed a significant temporal change and with log2 FC above 0.5 or below -0.5 **in the MARBLE cohort**. The row labels indicate the surgery type; the “Grand Mean” row represents data from all types of surgery combined. The values are represented as means (standard deviation).

|  | **FSTL3** | **SDF1** | **IBP3** | **SRGN** | **SAA1** |
| --- | --- | --- | --- | --- | --- |
| **Grand Mean** | 0.884 (0.033) | -0.776 (0.034) | 0.557 (0.029) | 0.809 (0.042) | 2.255 (0.122) |
| Thoracic | 0.77 (0.13) | -0.638 (0.133) | 0.482 (0.111) | 1.058 (0.166) | 2.408 (0.476) |
| General Surgery | 0.817 (0.087) | -0.715 (0.089) | 0.522 (0.074) | 0.911 (0.111) | 2.608 (0.319) |
| Gynecologic | 0.991 (0.147) | -0.723 (0.151) | 0.86 (0.126) | 0.697 (0.188) | 2.739 (0.54) |
| Orthopedic | 0.913 (0.054) | -0.791 (0.055) | 0.528 (0.046) | 0.808 (0.069) | 2.005 (0.198) |
| ENT Surgery | 1.04 (0.147) | -1.172 (0.151) | 0.557 (0.126) | 0.921 (0.188) | 2.18 (0.54) |
| Plastic Surgery | 1.069 (0.174) | -0.861 (0.179) | 0.808 (0.149) | 0.77 (0.222) | 1.304 (0.639) |
| Urologic | 0.832 (0.064) | -0.745 (0.066) | 0.544 (0.055) | 0.702 (0.082) | 2.429 (0.235) |
|  | **AGRG1** | **TSK** | **IBP1** | **PAI1** | **TIMP4** |
| **Grand Mean** | 0.773 (0.042) | 0.968 (0.06) | 0.99 (0.067) | 1.589 (0.108) | 0.714 (0.049) |
| Thoracic | 0.721 (0.167) | 0.768 (0.231) | 1.229 (0.257) | 1.112 (0.419) | 0.728 (0.191) |
| General Surgery | 0.739 (0.112) | 0.775 (0.155) | 1.372 (0.172) | 1.382 (0.281) | 0.593 (0.128) |
| Gynecologic | 0.702 (0.19) | 0.648 (0.262) | 0.701 (0.291) | 1.523 (0.475) | 0.876 (0.217) |
| Orthopedic | 0.789 (0.07) | 1.119 (0.096) | 0.798 (0.107) | 1.77 (0.174) | 0.705 (0.079) |
| ENT Surgery | 0.79 (0.19) | 1.503 (0.262) | 1.289 (0.291) | 1.793 (0.475) | 0.894 (0.217) |
| Plastic Surgery | 0.961 (0.224) | 0.91 (0.31) | 0.94 (0.345) | 2.603 (0.562) | 1.008 (0.256) |
| Urologic | 0.766 (0.083) | 0.874 (0.114) | 1 (0.127) | 1.398 (0.206) | 0.683 (0.094) |

* FSTL3: Follistatin-related protein 3; SDF1: Stromal cell-derived factor 1; IBP3: Insulin-like growth factor-binding protein 3; SRGN: Serglycin; SAA1: Serum amyloid A-1 protein; AGRG1: Adhesion G-protein coupled receptor G1; TSK: Tsukushin; IBP1: Insulin-like growth factor-binding protein 1; PAI1: Plasminogen activator inhibitor 1; TIMP4: Metalloproteinase inhibitor 4.

**Table S2a.** Proteins (gene names) used in the analysis of the temporal effect of pathway enrichment scores, for the up-regulated pathways **in the MARBLE cohort**.

| Pathway | Gene Names |
| --- | --- |
| negative regulation of smooth muscle cell migration | ABHD2, ADIPOQ, AIF1, BMP4, BMPR1A, CORO1B, GNA12, GNA13, IGFBP3, IGFBP5, LRP1, MEF2C, MIR1298, MIR137, MIR140, MIR15A, MIR182, MIR21, MIR214, MIR218-1, MIR223, MIR34A, MIR362, MIR424, MIR503, MIR638, MIR665, MYOCD, NDRG4, NF1, NFE2L2, PRKG1, SEMA6D, SERPINE1, SLIT2, TAFA5, TPM1, TRIB1 |
| negative regulation of endothelial cell apoptotic process | ABL1, ANGPT1, ANGPTL4, BRAF, CDH5, FGA, FGB, FGF21, FGG, GAS6, GATA2, GATA3, ICAM1, IL10, IL11, IL13, IL4, ITGB3, KDR, KRIT1, MAPK7, MIR126, MIR30B, MIR30E, MIR495, MIR590, NDNF, NFE2L2, PAK4, PDPK1, RAMP2, SCG2, SEMA5A, SERPINE1, TEK, TERT, TNFAIP3, TNIP2 |
| extrinsic apoptotic signaling pathway via death domain receptors | ARHGEF2, ATF3, BAD, BAG3, BAX, BCL2, BCL2L1, BEX3, BID, BLOC1S2, BMP5, BMPR1B, BRCA1, CASP10, CASP8, CASP8AP2, CFLAR, CRADD, DAB2IP, DAPK1, DAXX, DDX3X, DDX47, DEDD, DEDD2, DELE1, DIABLO, FADD, FAF1, FAIM, FAIM2, FASLG, FEM1B, FGA, FGB, FGG, GABARAP, GPX1, GSK3B, HGF, HMGB2, HMOX1, ICAM1, ITPRIP, LGALS3, MADD, MAL, MIR221, MIR222, MOAP1, NF1, NGF, NOS3, PARK7, PEA15, PIDD1, PIK3R1, PMAIP1, PTEN, RAF1, RFFL, RNF34, SCRT2, SERPINE1, SFRP1, SFRP2, SKIL, SORT1, SP100, SPI1, STK3, STK4, STX4, THBS1, TIMP3, TMBIM1, TMC8, TNF, TNFAIP3, TNFRSF10A, TNFRSF10B, TNFRSF10C, TNFRSF1A, TRADD, ZDHHC3, ZSWIM2 |
| regulation of extrinsic apoptotic signaling pathway via death domain receptors | ARHGEF2, ATF3, BCL2L1, BMP5, BMPR1B, BRCA1, CFLAR, DDX3X, FAF1, FAIM, FAIM2, FEM1B, FGA, FGB, FGG, GPX1, GSK3B, HGF, HMGB2, HMOX1, ICAM1, ITPRIP, LGALS3, MADD, MAL, MIR221, MIR222, NOS3, PARK7, PEA15, PMAIP1, PTEN, RAF1, RFFL, RNF34, SCRT2, SERPINE1, SFRP1, SFRP2, SKIL, SP100, STK3, STK4, STX4, THBS1, TIMP3, TMBIM1, TMC8, TNFAIP3, ZSWIM2 |
| smooth muscle cell migration | AAMP, ABHD2, ACE, ADAMTS1, ADIPOQ, AIF1, ATP7A, BCL2, BMP4, BMPR1A, CCL5, CCN3, CCN4, CORO1B, CRK, CYP1B1, DDIT3, DDR1, DDR2, DOCK4, DOCK5, DOCK7, FGF9, FOXO4, GNA12, GNA13, GRB10, HAS2, IGF1, IGFBP3, IGFBP5, ITGA2, ITGB3, LPAR1, LRP1, MAP3K7, MDK, MDM2, MEF2C, MIR1298, MIR135B, MIR137, MIR140, MIR143, MIR146A, MIR15A, MIR182, MIR20A, MIR21, MIR214, MIR218-1, MIR221, MIR223, MIR26A1, MIR302C, MIR34A, MIR362, MIR424, MIR448, MIR451A, MIR499A, MIR503, MIR638, MIR665, MYOCD, NDRG4, NF1, NFE2L2, NR4A3, NRP1, P2RY6, PARVA, PDGFA, PDGFB, PDGFD, PDGFRB, PLAT, PLAU, PLXNA1, POSTN, PRKG1, RPS6KB1, S100A11, SEMA6D, SERPINE1, SLIT2, SORL1, SRC, SSH1, TACR1, TAFA5, TERT, TLR4, TPM1, TRIB1, VTN |
| regulation of fibrinolysis | APOH, CPB2, F11, F12, F2, FAP, HRG, KLKB1, PLAT, PLAU, PLAUR, PLG, SERPINE1, SERPINF2, THBD, THBS1, USF1, VTN |
| muscle cell migration | AAMP, ABHD2, ACE, ADAMTS1, ADIPOQ, AIF1, AKIRIN1, ANXA1, ATP7A, BCL2, BIN3, BMP4, BMPR1A, CCL5, CCN3, CCN4, CORO1B, CRK, CYP1B1, DDIT3, DDR1, DDR2, DOCK4, DOCK5, DOCK7, FGF9, FOXO4, GNA12, GNA13, GRB10, HAS2, IGF1, IGFBP3, IGFBP5, ITGA2, ITGB1BP1, ITGB3, LPAR1, LRP1, MAP3K7, MDK, MDM2, MEF2C, MEGF10, MIR1298, MIR135B, MIR137, MIR140, MIR143, MIR146A, MIR15A, MIR182, MIR20A, MIR21, MIR214, MIR218-1, MIR221, MIR223, MIR26A1, MIR302C, MIR34A, MIR362, MIR424, MIR448, MIR451A, MIR499A, MIR503, MIR638, MIR665, MSTN, MYOCD, NDRG4, NET1, NF1, NFE2L2, NR4A3, NRP1, P2RY6, PARVA, PDGFA, PDGFB, PDGFD, PDGFRB, PLAT, PLAU, PLEKHO1, PLXNA1, POSTN, PRKG1, ROCK1, RPS6KB1, S100A11, SEMA6D, SERPINE1, SIX1, SIX4, SLIT2, SMO, SORL1, SRC, SSH1, TACR1, TAFA5, TCP11L2, TERT, THBS4, TLR4, TPM1, TRIB1, VTN |
| endothelial cell apoptotic process | ABL1, AKR1C3, ANGPT1, ANGPTL4, ANO6, BMPR2, BRAF, CCL2, CD160, CD248, CD40, CD40LG, CDH5, COL4A3, DAB2IP, ECSCR, FASLG, FGA, FGB, FGF21, FGG, GAS6, GATA2, GATA3, GPER1, HIPK1, HLA-G, ICAM1, IL10, IL11, IL13, IL4, ITGA4, ITGB3, KDR, KRIT1, MAP3K5, MAPK7, MIR101-1, MIR106B, MIR125A, MIR126, MIR132, MIR15A, MIR24-1, MIR30B, MIR30E, MIR375, MIR495, MIR590, NDNF, NFE2L2, PAK4, PDCD4, PDPK1, PLCG1, PRKCI, RAMP2, RGCC, SCG2, SEMA5A, SERPINE1, TEK, TERT, TGFB1, THBS1, TNF, TNFAIP3, TNIP2 |
| angiogenesis involved in wound healing | ADIPOR2, ALOX5, B4GALT1, CD34, CX3CL1, CXCR4, DAG1, FOXC2, GATA2, GPR4, GPX1, HPSE, ITGB3, KDR, MCAM, MIR1298, MIR200B, MIR34A, MIR451A, NDNF, NPR2, PIK3CB, PRCP, SERPINE1, SLC12A2, SMOC2, SRF, TAFA5, TNF, TNFAIP3, VEGFA, XBP1 |
| actin filament organization | ABI1, ABI2, ABITRAM, ABL1, ACTA1, ACTC1, ACTG1, ACTL8, ACTN1, ACTN2, ACTR2, ACTR3, ADD1, ADD2, ADD3, AIF1, AIF1L, ALDOA, ALMS1, ALOX15, ANG, AP1AR, APOA1, AQP2, ARAP1, ARF1, ARF6, ARFGEF1, ARFIP1, ARFIP2, ARHGAP12, ARHGAP17, ARHGAP18, ARHGAP25, ARHGAP28, ARHGAP35, ARHGAP40, ARHGAP6, ARHGEF10, ARHGEF10L, ARHGEF15, ARHGEF18, ARHGEF2, ARHGEF5, ARPC1A, ARPC1B, ARPC2, ARPC3, ARPC4, ARPC5, ARPC5L, ARPIN, ARRB1, ASAP3, AVIL, BAG4, BAIAP2, BAIAP2L1, BAIAP2L2, BBS4, BCAR1, BCL2, BIN1, BIN3, BRAF, BRK1, C15orf62, C9orf72, CALD1, CAP1, CAP2, CAPG, CAPZA1, CAPZA2, CAPZA3, CAPZB, CARMIL1, CARMIL2, CARMIL3, CATIP, CAV3, CCDC88A, CCL11, CCL21, CCL24, CCL26, CCN2, CCR7, CD2AP, CD47, CDC42, CDC42EP1, CDC42EP2, CDC42EP3, CDC42EP4, CDC42EP5, CFL1, CFL2, CGNL1, CLASP1, CLASP2, CLRN1, CNN1, CNN2, CNN3, COBL, CORO1A, CORO1B, CORO1C, CORO2B, CORO6, CORO7, COTL1, CRACD, CSF3, CTNNA2, CTTN, CUL3, CX3CL1, CXCL12, CYFIP1, CYFIP2, CYRIA, CYRIB, DAAM2, DBN1, DIAPH1, DIAPH2, DIAPH3, DLC1, DLG1, DMTN, DNAI3, DPYSL3, DSTN, ELMO1, ELMO2, ELMO3, ELN, EMP2, ENAH, EPHA1, EPS8, ERMN, ESAM, ESPN, ESPNL, EVL, EZR, F11R, F2RL1, FAM107A, FAM171A1, FAT1, FCHSD1, FCHSD2, FER, FERMT2, FHDC1, FHOD1, FHOD3, FLII, FLNA, FMN1, FMN2, FRMD7, FSCN1, FSCN2, FSCN3, GAS2, GAS2L1, GAS2L2, GAS2L3, GAS7, GBA2, GDPD2, GHRL, GHSR, GMFB, GMFG, GPR65, GRB2, GSN, HAX1, HCK, HCLS1, HIP1, HIP1R, HSP90B1, INF2, INPP5K, INPPL1, IQGAP1, IQGAP2, IQGAP3, ITGB1, ITGB1BP1, ITGB5, JAK2, JMY, KANK1, KANK2, KANK3, KANK4, KASH5, KBTBD13, KCTD13, KHDC3L, KIRREL1, KPTN, LATS1, LCP1, LIMA1, LIMCH1, LIMD2, LIMK1, LMOD1, LMOD2, LMOD3, LPAR1, MAD2L2, MAGEL2, MARCKS, MARCKSL1, MEGF9, MET, MICAL1, MICAL2, MICAL3, MICALL2, MIR138-1, MIR149, MIR20A, MIR21, MIR214, MKKS, MLST8, MSRB1, MSRB2, MTOR, MTPN, MTSS1, MYADM, MYO15A, MYO19, MYO1A, MYO1B, MYO1C, MYO1D, MYO1E, MYO1F, MYO1G, MYO1H, MYO5A, MYO5B, MYO5C, MYO6, MYO7A, MYO7B, MYOC, NAA80, NCK1, NCK2, NCKAP1, NCKAP1L, NEB, NEBL, NEDD9, NF2, NLRP5, NOSTRIN, NPHS1, NRAP, NRP1, OOEP, PACSIN1, PAK1, PAK2, PAK3, PAWR, PDCD10, PDLIM1, PDXP, PFN1, PFN2, PFN3, PFN4, PHACTR1, PHLDB2, PICK1, PIK3CA, PIK3R1, PIK3R2, PLA2G1B, PLEC, PLEK, PLEKHG2, PLEKHH2, PLS1, PLS3, POF1B, PPARGC1B, PPFIA1, PPM1E, PPM1F, PPP1R9A, PPP1R9B, PREX1, PRKCD, PRKCE, PRKCI, PRKN, PROX1, PSTPIP1, PSTPIP2, PTGER4, PTK2B, PXN, PYCARD, RAC1, RAC2, RAC3, RAPGEF3, RASA1, RDX, RFLNA, RFLNB, RGCC, RGS4, RHOA, RHOB, RHOBTB1, RHOBTB2, RHOC, RHOD, RHOF, RHOG, RHOH, RHOJ, RHOQ, RHOT1, RHOT2, RHOU, RHOV, RHPN1, RHPN2, RHPN2P1, RICTOR, RND1, RND2, RND3, RNH1, ROCK1, ROCK2, RUFY3, S100A10, S1PR1, SAMD14, SCIN, SDC4, SEMA5A, SERPINF2, SFRP1, SH3BP1, SH3D21, SH3KBP1, SHANK1, SHANK3, SHROOM1, SHROOM2, SHROOM3, SHROOM4, SHTN1, SLC9A1, SLIT2, SMAD3, SNX9, SORBS1, SORBS3, SPATC1L, SPIRE1, SPIRE2, SPTA1, SPTAN1, SPTB, SPTBN1, SPTBN2, SPTBN4, SPTBN5, SRC, SRF, SSH1, SSH2, SSH3, STMN1, SVIL, SWAP70, SYNPO, SYNPO2, SYNPO2L, TACR1, TACSTD2, TAGLN2, TAGLN3, TCAP, TENM1, TESK1, TF, TGFB3, TGFBR1, TJP1, TLE6, TMEFF2, TMOD1, TMOD2, TMOD3, TMOD4, TMSB10, TMSB15A, TMSB15B, TMSB15C, TMSB4X, TMSB4Y, TNFAIP1, TPM1, TPM2, TPM3, TPM4, TRIM27, TRIOBP, TRPV4, TSC1, TTC17, TTC8, TTN, TWF1, TWF2, USH1C, VASP, VIL1, VILL, WAS, WASF1, WASF2, WASF3, WASH3P, WASH6P, WASHC1, WASHC2A, WASHC2C, WASHC3, WASHC4, WASHC5, WASL, WDR1, WHAMM, WIPF1, WIPF3, WNT11, WNT4, XIRP1, XIRP2, ZBED3, ZEB2, ZYX |

**Table S2b.** Proteins (gene names) used in the analysis of the temporal effect of pathway enrichment scores, for down-regulated pathways **in the MARBLE cohort**.

| Pathway | Gene.names |
| --- | --- |
| regulation of non-canonical NF-kB signal transduction | ACTN4, ADGRG3, ADIPOR1, ADISSP, AGER, AGO3, AKIP1, APP, BCL3, BIRC2, BMP7, C1QTNF3, C1QTNF4, CALR, CCL19, CCN3, CD27, CD86, CPNE1, CYLD, DDX3X, DLG1, EDA, EDAR, EDN1, EGFR, EIF2AK2, EP300, EZR, GREM1, HAVCR2, HDAC7, IFI35, IL12B, IL18, IL18R1, IL1B, IL23A, ILK, IRAK1, LAPTM5, LGALS9, LIME1, LIMS1, LITAF, LRRC19, MIR132, MIR146A, MIR149, MIR15B, MIR182, MIR204, MIR21, MIR223, MIR27B, MIR29B1, MIR30C2, MIR508, MIR766, MIR9-1, MKRN2, NFAT5, NFKBIA, NLRC3, NLRP12, NLRP2, NLRP3, NMI, NOD1, NOD2, NR3C2, PDCD4, PHB1, PHB2, PPM1A, PPM1B, PRDX1, PTP4A3, PTPN22, PYCARD, PYDC2, RASSF2, RBCK1, RC3H1, RC3H2, RELA, RHOA, RIPK1, RPS3, RTKN2, SASH1, SPHK1, SPI1, TCIM, TERF2IP, TLR2, TLR3, TLR4, TLR7, TLR9, TMSB4X, TNF, TNFSF14, TRADD, TRAF6, TREM2, TRIM15, TRIM40, TRIM44, TRIM56, TRIM6, TRIM60, TRIP6, TSPAN6, UACA, ZC3H12A |
| negative regulation of leukocyte apoptotic process | ADA, ARG2, AURKB, AXL, BCL10, BCL11B, BCL2, BCL2L1, BCL3, BCL6, BMP4, CCL19, CCL21, CCL5, CCR5, CCR7, CD27, CD74, CXCL12, CXCR2, DOCK8, EFNA1, FADD, FOXP1, GAS6, GHSR, GPAM, HCLS1, HIF1A, HSH2D, IDO1, IL2, IL7R, IRF7, IRS2, ITPKB, JAK3, KIFAP3, KITLG, LILRB1, MERTK, MIF, MIR17HG, MIRLET7B, NOC2L, NOD2, ORMDL3, PDCD1, PIP, PRKCQ, PTCRA, RAG1, SELENOS, SLC39A10, SLC46A2, ST3GAL1, ST6GAL1, TSC22D3, VHL |
| sulfur compound metabolic process | AADAT, AASS, ABCB7, ABCD1, ABHD14B, ACACA, ACACB, ACAT1, ACLY, ACOT1, ACOT11, ACOT12, ACOT2, ACOT4, ACOT6, ACOT7, ACOT8, ACOT9, ACP3, ACSBG1, ACSBG2, ACSF2, ACSF3, ACSL1, ACSL3, ACSL4, ACSL5, ACSL6, ACSM1, ACSM2A, ACSM2B, ACSM3, ACSM4, ACSM5, ACSM6, ACSS1, ACSS2, ADI1, AGXT, AHCY, AHCYL1, AHCYL2, AKR1A1, ANGPT1, APIP, ARSG, B3GALT6, B3GAT3, B3GNT2, B3GNT3, B3GNT4, B3GNT7, B4GALT4, B4GAT1, BAAT, BHMT, BHMT2, BLMH, BOLA1, BOLA2, BOLA2B, BOLA3, BPNT1, BPNT2, BTD, CBS, CDO1, CHPF, CHPF2, CHST1, CHST11, CHST12, CHST13, CHST14, CHST15, CHST2, CHST3, CHST4, CHST5, CHST6, CHST7, CHST8, CHST9, CHSY1, CHSY3, CIAO1, CIAO2A, CIAO2B, CIAO3, CIAPIN1, CPS1, CSAD, CSGALNACT1, CSGALNACT2, CTH, CTNNB1, DGAT1, DGAT2, DIP2A, DLAT, DLD, DLST, DPEP1, DPEP2, DSE, DSEL, EDNRA, EDNRB, ELOVL1, ELOVL2, ELOVL3, ELOVL4, ELOVL5, ELOVL6, ELOVL7, ENOPH1, ENPP1, ENSG00000205794, ENSG00000274276, ETHE1, EXT1, EXT2, EXTL1, EXTL2, EXTL3, FAR1, FAR2, FDX2, FITM2, FMO1, FMO3, FXN, GAL3ST3, GAL3ST4, GCDH, GCLC, GCLM, GGT1, GGT2P, GGT3P, GGT5, GGT6, GGT7, GGTA1, GGTLC1, GGTLC2, GGTLC3, GHR, GLB1, GLCE, GLRX3, GLRX5, GLYAT, GNMT, GNS, GPAM, GPAT4, GPC1, GSTA1, GSTM1, GSTP1, GUSB, HACD1, HACD2, HEXA, HEXB, HGSNAT, HLCS, HMGCS1, HMGCS2, HPSE, HS2ST1, HS3ST1, HS3ST2, HS3ST3A1, HS3ST3B1, HS3ST4, HS3ST5, HS3ST6, HS6ST1, HS6ST2, HS6ST3, HSCB, HSD17B12, HSD17B4, HSPA9, HTD2, HYAL1, HYAL4, IBA57, ICMT, IDS, IDUA, ISCA1, ISCA2, ISCU, LIAS, LPO, LYRM4, MAT1A, MAT2A, MAT2B, MCCC1, MCEE, METTL16, MICAL1, MICAL2, MIR21, MLYCD, MMS19, MMUT, MPC2, MPST, MRI1, MSRA, MTAP, MTHFD1, MTHFD2L, MTHFR, MTR, MTRR, MVD, MVK, NAGLU, NDNF, NDOR1, NDST1, NDST2, NDST3, NDST4, NDUFAB1, NFS1, NFU1, NOX4, NUBP1, NUBP2, NUBPL, NUDT19, NUDT7, NUDT8, OGDH, OXSM, PAPSS1, PAPSS2, PAX8, PDHA1, PDHA2, PDHB, PDHX, PDK1, PDK2, PDK3, PDK4, PHGDH, PIPOX, PMVK, PPCS, PPT1, PPT2, PXYLP1, SGSH, SLC10A7, SLC19A2, SLC19A3, SLC25A1, SLC25A10, SLC25A19, SLC27A2, SLC35B2, SLC35D2, SLC5A6, SLC7A11, SMS, SNCA, SP1, SQOR, ST3GAL1, ST3GAL2, ST3GAL3, ST3GAL4, ST3GAL6, STAT5A, STAT5B, SUCLA2, SUCLG1, SUCLG2, SULT1A1, SULT1A2, SULT1A3, SULT1A4, SULT1B1, SULT1C2, SULT1C3, SULT1C4, SULT1E1, SULT2A1, SULT2B1, SULT4A1, SULT6B1, SUOX, TCF7L2, TDO2, TECR, THEM5, THTPA, TKTL1, TM9SF2, TPK1, TPST1, TPST2, TST, TSTD1, UGDH, UST, XDH, XYLT1, XYLT2 |
| proteoglycan metabolic process | ADAMTS12, ADAMTS7, B3GALT6, B3GAT1, B3GAT2, B3GAT3, B4GALT7, BMP2, BMPR1B, BMPR2, BPNT2, BTK, CANT1, CHPF, CHPF2, CHST10, CHST11, CHST12, CHST13, CHST14, CHST15, CHST3, CHST7, CHST8, CHST9, CHSY1, CHSY3, CNMD, COL11A1, COL2A1, CSGALNACT1, CSGALNACT2, CTNNB1, CYTL1, DSE, DSEL, EXT1, EXT2, EXTL1, EXTL2, EXTL3, FAM20B, FOXL1, GAL3ST3, GAL3ST4, GLB1, GLCE, GPC1, GUSB, HEXA, HEXB, HGSNAT, HPSE, HS2ST1, HS3ST1, HS3ST2, HS3ST3A1, HS3ST3B1, HS3ST4, HS3ST5, HS3ST6, HS6ST1, HS6ST2, HS6ST3, HYAL1, HYAL4, IDS, IDUA, IGF1, IHH, MUSTN1, NAGLU, NDNF, NDST1, NDST2, NDST3, NDST4, PXYLP1, SGSH, SLC2A10, SLC35B2, SLC35D2, SULF1, SULF2, TCF7L2, TM9SF2, UGDH, UST, XYLT1, XYLT2 |
| leukocyte apoptotic process | ADA, ADAM17, ADAM8, AKT1, ANXA1, ARG2, AURKB, AXL, BAK1, BAX, BCL10, BCL11B, BCL2, BCL2L1, BCL2L11, BCL3, BCL6, BIRC7, BMP4, BTK, CASP3, CASP7, CASP9, CCL19, CCL21, CCL5, CCR5, CCR7, CD27, CD274, CD3G, CD74, CDKN2A, CHEK2, CLC, CRKL, CTSL, CXCL12, CXCR2, DFFA, DNAJA3, DOCK8, EBF4, EFNA1, FADD, FAS, FASLG, FNIP1, FOXP1, GAS6, GHSR, GIMAP8, GLI3, GPAM, HCAR2, HCLS1, HIF1A, HSH2D, IDO1, IL10, IL2, IL2RA, IL6, IL7R, IRF3, IRF7, IRS2, ITPKB, JAK3, KDELR1, KIFAP3, KITLG, LGALS16, LGALS3, LGALS9, LILRB1, LIPA, LMBR1L, LYN, MEF2C, MERTK, MIAT, MIF, MIR17HG, MIR34A, MIRLET7B, NF1, NFKBIZ, NOC2L, NOD2, ORMDL3, P2RX7, PDCD1, PIK3CB, PIK3CD, PIP, PKN1, PLEKHO2, PRELID1, PRKCQ, PRKD2, PTCRA, RAG1, RAPGEF2, RIPK1, RIPK3, RPS6, SELENOS, SIRT1, SLC39A10, SLC46A2, SLC7A11, ST3GAL1, ST6GAL1, TNFRSF21, TP53, TRAF3IP2, TSC22D3, VHL, WNT5A, ZC3H8 |
| neuron projection extension involved in neuron projection guidance | ALCAM, BMPR2, CXCL12, DSCAM, HDAC6, MEGF8, NRP1, NRP2, PLXNA3, PLXNA4, RYK, SEMA3A, SEMA3B, SEMA3C, SEMA3D, SEMA3E, SEMA3F, SEMA3G, SEMA4A, SEMA4B, SEMA4C, SEMA4D, SEMA4F, SEMA4G, SEMA5A, SEMA5B, SEMA6A, SEMA6B, SEMA6C, SEMA6D, SEMA7A, SLIT1, SLIT2, SLIT3, VEGFA, WNT3, WNT3A, WNT5A |
| regulation of leukocyte apoptotic process | ADA, ADAM17, ADAM8, ANXA1, ARG2, AURKB, AXL, BAX, BCL10, BCL11B, BCL2, BCL2L1, BCL2L11, BCL3, BCL6, BIRC7, BMP4, BTK, CCL19, CCL21, CCL5, CCR5, CCR7, CD27, CD274, CD3G, CD74, CDKN2A, CXCL12, CXCR2, DOCK8, EFNA1, FADD, FNIP1, FOXP1, GAS6, GHSR, GIMAP8, GPAM, HCAR2, HCLS1, HIF1A, HSH2D, IDO1, IL10, IL2, IL7R, IRF7, IRS2, ITPKB, JAK3, KIFAP3, KITLG, LGALS16, LGALS3, LGALS9, LILRB1, LYN, MEF2C, MERTK, MIAT, MIF, MIR17HG, MIR34A, MIRLET7B, NF1, NOC2L, NOD2, ORMDL3, P2RX7, PDCD1, PIK3CB, PIK3CD, PIP, PRELID1, PRKCQ, PRKD2, PTCRA, RAG1, RAPGEF2, RIPK3, SELENOS, SIRT1, SLC39A10, SLC46A2, SLC7A11, ST3GAL1, ST6GAL1, TP53, TSC22D3, VHL, WNT5A, ZC3H8 |
| response to chemokine | ACKR1, ACKR2, ACKR3, ACKR4, CCL1, CCL11, CCL13, CCL14, CCL15, CCL16, CCL17, CCL18, CCL19, CCL2, CCL20, CCL21, CCL22, CCL23, CCL24, CCL25, CCL26, CCL3, CCL3L1, CCL3L3, CCL4, CCL5, CCL7, CCL8, CCR1, CCR10, CCR2, CCR3, CCR4, CCR5, CCR6, CCR7, CCR8, CCR9, CCRL2, CIB1, CMKLR1, CX3CL1, CX3CR1, CXCL1, CXCL10, CXCL11, CXCL12, CXCL13, CXCL2, CXCL3, CXCL5, CXCL6, CXCL8, CXCL9, CXCR1, CXCR2, CXCR3, CXCR4, CXCR5, CXCR6, DOCK8, DUSP1, EDN1, ENTREP1, FOXC1, GPR17, GPR35, GPR75, HIF1A, LOX, LRCH1, LYN, MPL, OXSR1, PADI2, PF4, PF4V1, PPBP, PTK2B, RBM15, RHOA, RIPOR2, RNF113A, ROBO1, SH2B3, SLC12A2, SLIT2, SLIT3, STK39, TFF2, THPO, TREM2, WBP1L, WNK1, XCL1, XCL2, XCR1, ZC3H12A |
| glycoprotein metabolic process | A4GALT, A4GNT, AATF, ABCA2, ABCA7, ACER2, ACOT8, ADAMTS12, ADAMTS13, ADAMTS7, AGA, AGO2, ALG1, ALG10, ALG10B, ALG11, ALG12, ALG13, ALG14, ALG1L2, ALG2, ALG3, ALG5, ALG6, ALG8, ALG9, APCS, AQP11, ARFGEF1, ATP7A, B3GALNT1, B3GALNT2, B3GALT1, B3GALT2, B3GALT4, B3GALT5, B3GALT6, B3GALT9, B3GAT1, B3GAT2, B3GAT3, B3GLCT, B3GNT2, B3GNT3, B3GNT4, B3GNT5, B3GNT6, B3GNT7, B3GNT8, B3GNT9, B4GALNT2, B4GALT1, B4GALT5, B4GALT7, B4GAT1, BACE2, BCL2, BMP2, BMPR1B, BMPR2, BPNT2, BTK, C1GALT1, C1GALT1C1, C1GALT1C1L, C20orf173, CANT1, CCL19, CCL21, CCR7, CELA1, CHP1, CHPF, CHPF2, CHST10, CHST11, CHST12, CHST13, CHST14, CHST15, CHST3, CHST4, CHST7, CHST8, CHST9, CHSY1, CHSY3, CNMD, COG3, COG7, COL11A1, COL2A1, CRPPA, CSGALNACT1, CSGALNACT2, CST3, CTNNB1, CTSL, CYTL1, DAD1, DDOST, DERL3, DHDDS, DOLK, DOLPP1, DPAGT1, DPM1, DPM2, DPM3, DPY19L1, DPY19L2, DPY19L2P2, DPY19L3, DPY19L4, DSE, DSEL, EDEM1, EDEM2, EDEM3, ENGASE, ENTPD5, EOGT, ERP44, EXT1, EXT2, EXTL1, EXTL2, EXTL3, FAM20B, FBXO17, FBXO2, FBXO27, FBXO44, FBXO6, FKRP, FKTN, FOXL1, FUT1, FUT10, FUT2, FUT3, FUT4, FUT5, FUT6, FUT7, FUT8, FUT9, GAL3ST1, GAL3ST2, GAL3ST3, GAL3ST4, GALNT1, GALNT10, GALNT11, GALNT12, GALNT13, GALNT14, GALNT15, GALNT16, GALNT17, GALNT18, GALNT2, GALNT3, GALNT4, GALNT5, GALNT6, GALNT7, GALNT8, GALNT9, GALNTL5, GALNTL6, GANAB, GANC, GATA1, GCNT1, GCNT2, GCNT3, GCNT4, GFPT1, GFPT2, GLB1, GLCE, GMPPB, GNPTAB, GNPTG, GOLGA2, GOLPH3, GPC1, GUSB, GXYLT1, GXYLT2, HBEGF, HEXA, HEXB, HGSNAT, HIF1A, HPSE, HS2ST1, HS3ST1, HS3ST2, HS3ST3A1, HS3ST3B1, HS3ST4, HS3ST5, HS3ST6, HS6ST1, HS6ST2, HS6ST3, HYAL1, HYAL4, IDS, IDUA, IGF1, IHH, IL15, IL33, ITM2A, ITM2B, ITM2C, JAK3, KRTCAP2, LARGE1, LARGE2, LEP, MAGT1, MAN1A1, MAN1A2, MAN1B1, MAN1C1, MAN2A1, MAN2A2, MAN2B1, MANBA, MGAT1, MGAT2, MGAT3, MGAT4A, MGAT4B, MGAT4C, MGAT4D, MGAT5, MGAT5B, MIR101-1, MIR106A, MIR144, MIR147A, MIR153-1, MIR17, MIR181B1, MIR20A, MIR298, MIR31, MIR323A, MIR455, MIR520C, MIR644A, MLEC, MMP12, MOGS, MPDU1, MUSTN1, NAGLU, NCCRP1, NCSTN, NDNF, NDST1, NDST2, NDST3, NDST4, NECAB1, NECAB2, NECAB3, NEU2, NEU4, NGLY1, NPC1, NUDT14, NUS1, OGA, OGT, OST4, OSTC, PARK7, PAWR, PCSK6, PGM3, PHLDA1, PLCB1, PLOD1, PLOD2, PLOD3, PMM1, PMM2, POFUT1, POFUT2, POGLUT1, POGLUT2, POGLUT3, POMGNT1, POMGNT2, POMK, POMT1, POMT2, PORCN, PRKCSH, PTX3, PXYLP1, RAB1A, RAB1B, RAMP1, RFT1, RPN1, RPN2, RXYLT1, SERP1, SGSH, SLC2A10, SLC34A1, SLC35B2, SLC35C1, SLC35D2, SLC39A8, SLC51B, SOAT1, SRD5A3, ST3GAL1, ST3GAL2, ST3GAL3, ST3GAL4, ST3GAL5, ST3GAL6, ST6GAL1, ST6GALNAC1, ST6GALNAC2, ST6GALNAC3, ST6GALNAC4, ST6GALNAC6, ST8SIA1, ST8SIA2, ST8SIA3, ST8SIA4, ST8SIA5, ST8SIA6, STT3A, STT3B, SULF1, SULF2, TCF7L2, TET1, TET2, TET3, TM9SF2, TMEM106A, TMEM165, TMEM258, TMEM59, TMTC1, TMTC2, TMTC3, TMTC4, TNIP1, TRAK1, TREX1, TUSC3, UBE2J1, UGDH, UGGT1, UGGT2, UST, XXYLT1, XYLT1, XYLT2 |
| negative regulation of axon extension involved in axon guidance | HDAC6, NRP1, PLXNA3, RYK, SEMA3A, SEMA3B, SEMA3C, SEMA3D, SEMA3E, SEMA3F, SEMA3G, SEMA4A, SEMA4B, SEMA4C, SEMA4D, SEMA4F, SEMA4G, SEMA5A, SEMA5B, SEMA6A, SEMA6B, SEMA6C, SEMA6D, SEMA7A, SLIT1, WNT3, WNT3A, WNT5A |

**ApoE-e4 effect on protein intensity levels, and changes over time, in the MARBLE Cohort**

Next, we conducted a sensitivity analysis to assess the effect of *APOE4* carrier status on CSF protein levels in the MARBLE cohort. After FDR adjustment, there was no significant effect of *APOE4* carrier status, and no significant interaction effect of *APOE4* carrier status by time, on CSF protein levels. This demonstrates that, after multiple comparison correction, *APOE4* carriers did not have significant differences in the levels of any CSF protein, or in the changes in these CSF protein levels from before to 24 hours after surgery.

See Supplemental Excel file – Sheet 5. The color-coded columns (beta_time, beta_e4, and beta_interaction) refer to the beta coefficients of time, *APOE4* carrier status, and interaction between time and *APOE4* carrier status. The red color indicates more positive effect (i.e., up-regulation), and the blue color indicates more negative effect (i.e., down-regulation). Proteins with significant adjusted p-values are coded with borders.

Among a total of 2086 proteins, there were 769 proteins that showed a significant time effect with FDR adjusted p-value<0.05 (390 positive effect; 379 negative effect), after adjusting for *APOE4* carrier status and interaction between time and *APOE4* carrier status. There were no proteins that showed a significant main effect of *APOE4* carrier status, and no significant interaction effect between *APOE4* carrier status and time on preoperative to 24 hr protein level changes.

**CN-105 Effect on Postoperative Protein and Pathway Changes in the MARBLE Trial Patients**

### **Table S3**. Baseline characteristics of the 137 participants from the **MARBLE TRIAL*** separating the placebo and CN-105 groups.

|  | Placebo (37, 27.0%) | CN-105** (100, 73.0%) |
| --- | --- | --- |
| **Age** | 69.1 (5.0) | 68.9 (5.1) |
| **Sex** |  |  |
| Male | 24 (64.9%) | 65 (65%) |
| Female | 13 (35.1%) | 35 (35%) |
| **Education (years)** | 16.2 (2.9) | 16.4 (3.6) |
| **Race** |  |  |
| Caucasian/White | 31 (83.8%) | 87 (87%) |
| Black or African American | 4 (10.8%) | 6 (6%) |
| Asian | 0 (0%) | 2 (2%) |
| American Indian or Alaska Native | 0 (0%) | 3 (3%) |
| Other | 2 (5.4%) | 2 (2%) |
| **ApoE-e2 copy number** | |  |
| 0 | 36 (97.3%) | 86 (86%) |
| 1 | 1 (2.7%) | 13 (13%) |
| 2 | 0 (0%) | 1 (1%) |
| **ApoE-e4 copy number**** | |  |
| 0 | 26 (70.3%) | 73 (73%) |
| 1 | 10 (27.0%) | 25 (25%) |
| 2 | 1 (2.7%) | 2 (2%) |
| **BMI** | 29.3 (3.7) | 29.8 (5.5) |
| **Surgery type** | |  |
| Thoracic Surgery | 2 (5.4%) | 7 (7%) |
| General Surgery | 3 (8.1%) | 17 (17%) |
| Gynecologic Surgery | 4 (10.8%) | 3 (3%) |
| Orthopedics Surgery | 13 (35.1%) | 39 (39%) |
| ENT Surgery | 3 (8.1%) | 4 (4%) |
| Plastic Surgery | 1 (2.7%) | 4 (4%) |
| Urologic Surgery | 11 (29.7%) | 26 (26%) |
| **ASA physical status classification** | |  |
| 1 | 0 (0%) | 1 (1%) |
| 2 | 14 (37.8%) | 33 (33%) |
| 3 | 22 (59.5%) | 66 (66%) |
| 4 | 1 (2.7%) | 0 (0%) |

*See Figure S1 for CONSORT diagram of MARBLE trial enrollment.

**The CN-105 group includes 38 participants who received 0.1mg/kg, 29 who received 0.5 mg/kg, and 33 who received 1.0 mg/kg of CN-105.

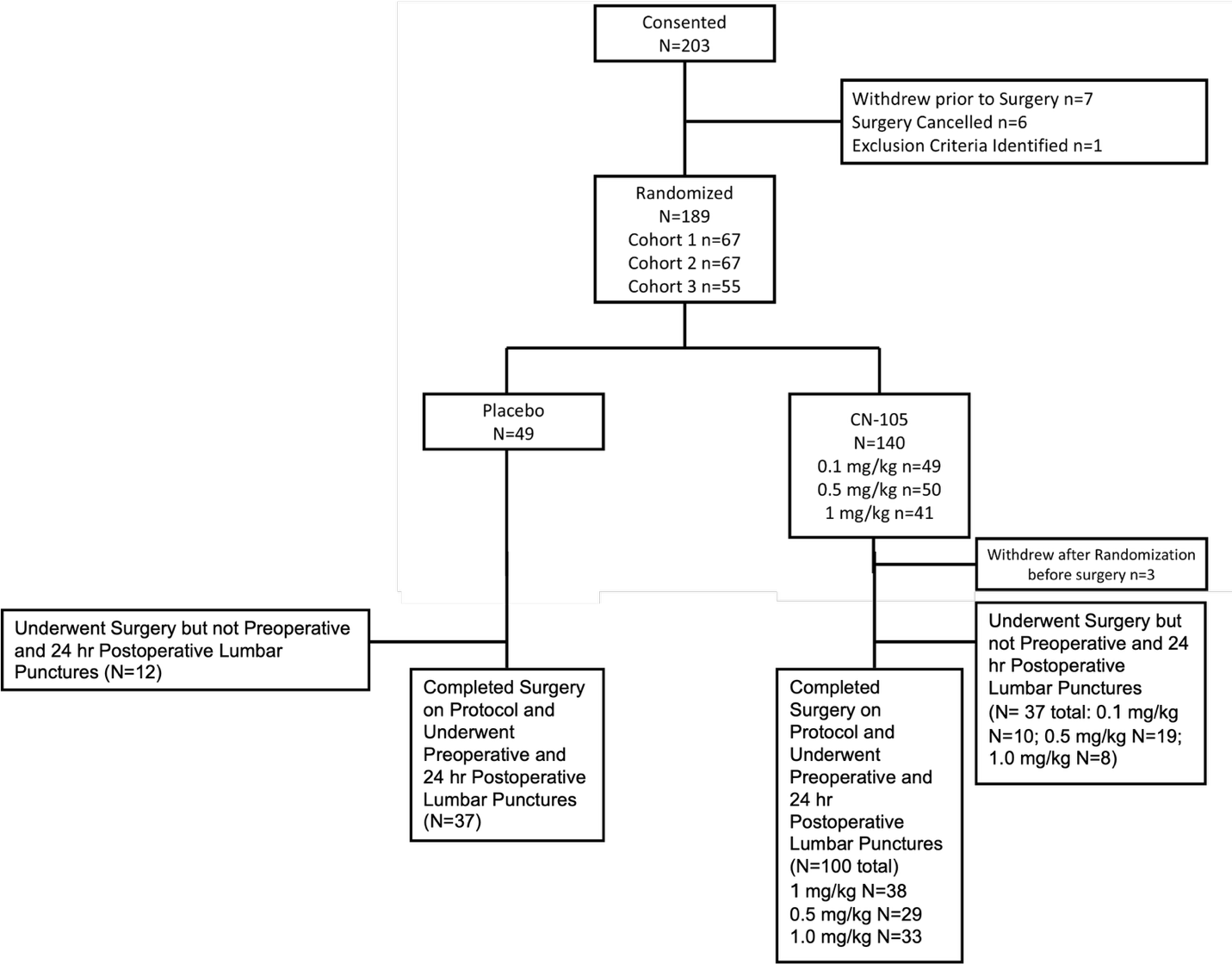

**Figure S1.** CONSORT diagram for the MARBLE trial.

**Table S4.** Univariable effect of CN-105 (vs placebo) on postoperative CSF protein level changes (10 up-regulated and 10 down-regulated, sorted by p-value) from **the MARBLE cohort**. Positive Log_2_ fold change (Log_2_ FC) indicate a positive effect of CN-105, and *vice versa*.

| **Up-regulated proteins** | | **Log_2_ FC** | **Std.Err** | **t** | **p-value** | **Adj.P** |
| --- | --- | --- | --- | --- | --- | --- |
| DNJC3 | DnaJ Heat Shock Protein Family (Hsp40) Member C3 | 0.095 | 0.030 | 3.157 | 1.97E-03 | 0.734 |
| ABCA2 | ATP Binding Cassette Subfamily A Member 2 | 0.130 | 0.042 | 3.090 | 2.43E-03 | 0.734 |
| DPP6 | Dipeptidyl Peptidase Like 6 | 0.194 | 0.065 | 2.991 | 3.30E-03 | 0.734 |
| S38AA | Solute Carrier Family 38 Member 10 | 0.118 | 0.041 | 2.907 | 4.26E-03 | 0.734 |
| T132E | Transmembrane Protein 132E | 0.219 | 0.077 | 2.849 | 5.07E-03 | 0.734 |
| K319L | KIAA0319 Like | 0.085 | 0.030 | 2.795 | 5.95E-03 | 0.734 |
| KAT6B | Lysine Acetyltransferase 6B | 0.636 | 0.235 | 2.703 | 7.76E-03 | 0.734 |
| PPIB | Peptidylprolyl Isomerase B | 0.062 | 0.023 | 2.645 | 9.13E-03 | 0.734 |
| MERTK | MER Proto-Oncogene, Tyrosine Kinase | 0.214 | 0.081 | 2.644 | 9.15E-03 | 0.734 |
| LEP | Leptin | 0.499 | 0.190 | 2.627 | 9.61E-03 | 0.734 |
| **Down-regulated proteins** | | **Log_2_ FC** | **Std.Err** | **t** | **p-value** | **Adj.P** |
| AK1A1 | Aldo-Keto Reductase Family 1 Member A1 | -0.451 | 0.108 | -4.179 | 5.23E-05 | 0.109 |
| SCUB1 | Signal Peptide, CUB Domain And EGF Like Domain Containing 1 | -0.192 | 0.056 | -3.423 | 8.22E-04 | 0.577 |
| MEGF6 | Multiple EGF Like Domains 6 | -0.519 | 0.152 | -3.420 | 8.30E-04 | 0.577 |
| PPIA | Peptidylprolyl Isomerase A | -0.161 | 0.051 | -3.136 | 2.10E-03 | 0.734 |
| TNR14 | TNF Receptor Superfamily Member 14 | -0.253 | 0.086 | -2.936 | 3.91E-03 | 0.734 |
| SRGN | Serglycin | -0.271 | 0.093 | -2.914 | 4.18E-03 | 0.734 |
| SAA1 | Serum Amyloid A1 | -0.778 | 0.268 | -2.901 | 4.34E-03 | 0.734 |
| FBN1 | Fibrillin 1 | -0.066 | 0.023 | -2.882 | 4.60E-03 | 0.734 |
| CLIC1 | Chloride Intracellular Channel 1 | -0.201 | 0.070 | -2.853 | 5.01E-03 | 0.734 |
| DSG2 | Desmoglein 2 | -0.081 | 0.029 | -2.826 | 5.43E-03 | 0.734 |

* Abbreviations: Std.Err: standard error; Adj.P: adjusted p-value. FC: fold change.

**Table S5.** The 10 proteins (sorted by p-values, smallest to largest) from the linear models that assessed the effect of CN-105 **dose level** (0 mg/kg, 0.1 mg/kg, 0.5 mg/kg, 1.0 mg/kg) on CSF protein intensity change from before to 24 hours after surgery **in the MARBLE cohort**. The linear model included the dose level as a continuous variable (0, 0.1, 0.5, or 1 mg/kg).

| **Up-regulated proteins** | | **Log_2_ FC** | **Std.Err** | **t** | **p-value** | **Adj.P** |
| --- | --- | --- | --- | --- | --- | --- |
| LRRC4 | Leucine-rich repeat-containing protein 4 | 0.438 | 0.123 | 3.562 | 5.09E-04 | 0.212 |
| PPIC | Peptidyl-prolyl cis-trans isomerase C | 0.184 | 0.058 | 3.185 | 1.80E-03 | 0.369 |
| GALT7 | N-acetylgalactosaminyltransferase 7 | 0.108 | 0.034 | 3.178 | 1.84E-03 | 0.369 |
| PCDGL | Protocadherin gamma-C4 | 0.536 | 0.171 | 3.133 | 2.12E-03 | 0.369 |
| CNTP3 | Contactin-associated protein-like 3 | 0.143 | 0.046 | 3.098 | 2.37E-03 | 0.381 |
| DPP6 | Dipeptidyl aminopeptidase-like protein 6 | 0.219 | 0.073 | 3.014 | 3.08E-03 | 0.460 |
| DEN1B | DENN domain-containing protein 1B | 0.263 | 0.088 | 2.978 | 3.44E-03 | 0.478 |
| TMED9 | Transmembrane emp24 domain-containing protein 9 | 0.370 | 0.126 | 2.928 | 4.00E-03 | 0.491 |
| KCC2B | Calcium/calmodulin-dependent protein kinase type II subunit beta | 0.125 | 0.043 | 2.897 | 4.40E-03 | 0.510 |
| LEP | Leptin | 0.607 | 0.212 | 2.863 | 4.87E-03 | 0.535 |
| **Down-regulated proteins** | | **Log_2_ FC** | **Std.Err** | **t** | **p-value** | **Adj.P** |
| HEPC | Hepcidin | -1.005 | 0.265 | -3.799 | 2.19E-04 | 0.176 |
| TNR14 | Tumor necrosis factor receptor superfamily member 14 | -0.357 | 0.095 | -3.763 | 2.50E-04 | 0.176 |
| ZN184 | Zinc finger protein 184 | -1.123 | 0.299 | -3.759 | 2.53E-04 | 0.176 |
| ILRL1 | Interleukin-1 receptor-like 1 | -1.755 | 0.489 | -3.591 | 4.61E-04 | 0.212 |
| SCUB1 | Signal peptide, CUB and EGF-like domain-containing protein 1 | -0.201 | 0.063 | -3.180 | 1.83E-03 | 0.369 |
| DSC1 | Desmocollin-1 | -0.411 | 0.131 | -3.147 | 2.03E-03 | 0.369 |
| LTBP4 | Latent-transforming growth factor beta-binding protein 4 | -0.088 | 0.028 | -3.138 | 2.09E-03 | 0.369 |
| TRY2 | Trypsin-2 | -0.692 | 0.221 | -3.137 | 2.09E-03 | 0.369 |
| FGL2 | Fibroleukin | -0.129 | 0.044 | -2.932 | 3.95E-03 | 0.491 |
| SBSPO | Somatomedin-B and thrombospondin type-1 domain-containing protein | -0.201 | 0.072 | -2.768 | 6.43E-03 | 0.612 |

* The linear models included covariates: CN-105 dose as a continuous variable (0, 0.1, 0.5, or 1 mg/kg).

* Abbreviations: Adj.P: adjusted p-value.

**Table S6.** Pathways (10 up-regulated and 10 down-regulated, sorted by p-value) that show differential pre to 24-hr post-op change among CN-105 vs placebo treated patients, based on the Gene Ontology database for biological processes (GO-BP) **in the MARBLE cohort**. The univariable linear models were fit for the 1001 pathways that showed a significant temporal effect. The top panel displays the pathways that are up-regulated with CN-105. The bottom panel displays the pathways that are down-regulated with CN-105. Beta coefficients are from linear models that assessed the effect of CN-105 on the ssGSEA enrichment scores.

| **Up-regulated pathways** | **Beta** | **Std.Err** | **t** | **p-value** | **Adj.P** |
| --- | --- | --- | --- | --- | --- |
| negative regulation of secretion | 24.876 | 7.031 | 3.538 | 5.53E-04 | 0.397 |
| leukocyte tethering or rolling | 43.800 | 12.758 | 3.433 | 7.92E-04 | 0.397 |
| hormone transport | 17.383 | 5.716 | 3.041 | 2.83E-03 | 0.731 |
| glycoprotein metabolic process | 20.673 | 6.908 | 2.993 | 3.29E-03 | 0.731 |
| cytokine production involved in inflammatory response | 31.337 | 10.667 | 2.938 | 3.89E-03 | 0.731 |
| oligosaccharide metabolic process | 24.797 | 8.806 | 2.816 | 5.59E-03 | 0.731 |
| insulin receptor signaling pathway | 30.390 | 10.810 | 2.811 | 5.67E-03 | 0.731 |
| glycoprotein biosynthetic process | 20.347 | 7.283 | 2.794 | 5.97E-03 | 0.731 |
| n glycan processing | 41.575 | 15.060 | 2.761 | 6.57E-03 | 0.731 |
| proteoglycan biosynthetic process | 23.476 | 8.721 | 2.692 | 8.00E-03 | 0.801 |
| **Down-regulated pathways** | **Beta** | **Std.Err** | **t** | **p-value** | **Adj.P** |
| regulation of muscle contraction | -26.164 | 10.495 | -2.493 | 1.39E-02 | 0.882 |
| regulation of protein dephosphorylation | -33.520 | 13.581 | -2.468 | 1.48E-02 | 0.882 |
| regulation of striated muscle contraction | -30.342 | 12.537 | -2.420 | 1.68E-02 | 0.882 |
| regulation of cardiac muscle contraction | -33.807 | 14.067 | -2.403 | 1.76E-02 | 0.882 |
| regulation of protein localization to cell periphery | -18.277 | 8.154 | -2.242 | 2.66E-02 | 0.897 |
| regulation of actin filament based process | -12.712 | 5.673 | -2.241 | 2.67E-02 | 0.897 |
| primary alcohol metabolic process | -30.936 | 13.855 | -2.233 | 2.72E-02 | 0.897 |
| actin polymerization or depolymerization | -19.689 | 8.918 | -2.208 | 2.90E-02 | 0.897 |
| regulation of calcium ion transmembrane transporter activity | -32.527 | 14.935 | -2.178 | 3.12E-02 | 0.897 |
| terpenoid metabolic process | -31.866 | 14.878 | -2.142 | 3.40E-02 | 0.897 |

* Abbreviations: Std.Err: standard error; Adj.P: adjusted p-value.

* See Supplemental Digital Content Table S10 for multivariable model adjusted for CN-105, age, sex, education years, *ApoE*-e2 copy number, *ApoE*-e4 copy number, BMI, surgery type, ASA physical status classification, and baseline enrichment score.

**Multivariable analysis of protein intensity levels and pathway enrichment scores, and modulation by CN-105, in the MARBLE Cohort**

Continuous covariates (age, years of education, BMI, baseline protein/pathway intensities) were standardized to have a mean 0 and variance 1.

**Table S7**. The top 10 p-values from multivariable linear models that assessed the temporal changes in protein levels from before to 24 hours after surgery **in the MARBLE cohort**. The linear models were adjusted for CN-105 (all dose groups vs placebo), age, sex (male as the reference group), years of education, ApoE-e2 copy number (0, 1, or 2 alleles), ApoE-e4 copy number (0, 1, or 2 alleles), BMI, surgery type (thoracic vs others), ASA physical status classification (1 or 2 as the reference group), and baseline protein intensity. Positive Log_2_ fold change (Log_2_ FC) coefficients indicate an increase in the given protein level from before to 24 hours after surgery, and *vice versa*.

| **Up-regulated proteins** | | **Log_2_ FC** | **Std.Err** | **t** | **p-value** | **Adj.P** |
| --- | --- | --- | --- | --- | --- | --- |
| NID1 | Nidogen 1 | 0.527 | 0.039 | 13.626 | <0.001 | <0.001 |
| FSTL3 | Follistatin Like 3 | 1.012 | 0.076 | 13.229 | <0.001 | <0.001 |
| SRGN | Serglycin | 1.009 | 0.098 | 10.335 | <0.001 | <0.001 |
| SAA1 | Serum Amyloid A1 | 2.611 | 0.259 | 10.082 | <0.001 | <0.001 |
| PAI1 | Serpin Family E Member 1 | 1.796 | 0.187 | 9.622 | <0.001 | <0.001 |
| OMD | Osteomodulin | 0.413 | 0.043 | 9.500 | <0.001 | <0.001 |
| TSK | IL2 Inducible T Cell Kinase | 1.256 | 0.133 | 9.430 | <0.001 | <0.001 |
| MMRN1 | Multimerin 1 | 0.496 | 0.054 | 9.152 | <0.001 | <0.001 |
| IBP3 | Insulin Like Growth Factor Binding Protein 3 | 0.591 | 0.067 | 8.876 | <0.001 | <0.001 |
| CCN2 | Cellular Communication Network Factor 2 | 0.505 | 0.058 | 8.734 | <0.001 | <0.001 |
| **Down-regulated proteins** | | **Log_2_ FC** | **Std.Err** | **t** | **p-value** | **Adj.P** |
| SDF1 | Stromal Cell-Derived Factor 1 | -0.909 | 0.089 | -10.236 | <0.001 | <0.001 |
| ITIH5 | Inter-Alpha-Trypsin Inhibitor Heavy Chain 5 | -0.288 | 0.028 | -10.149 | <0.001 | <0.001 |
| CO1A1 | Collagen Type I Alpha 1 Chain | -0.354 | 0.038 | -9.361 | <0.001 | <0.001 |
| CO1A2 | Collagen Type I Alpha 2 Chain | -0.312 | 0.037 | -8.402 | <0.001 | <0.001 |
| PPIB | Peptidylprolyl Isomerase B | -0.219 | 0.027 | -8.232 | <0.001 | <0.001 |
| CO3A1 | Collagen Type III Alpha 1 Chain | -0.278 | 0.036 | -7.670 | <0.001 | <0.001 |
| ADA2 | Adenosine Deaminase 2 | -0.423 | 0.055 | -7.669 | <0.001 | <0.001 |
| CBPQ | Carboxypeptidase Q | -0.176 | 0.023 | -7.592 | <0.001 | <0.001 |
| LCAT | Lecithin-Cholesterol Acyltransferase | -0.206 | 0.027 | -7.560 | <0.001 | <0.001 |
| SPON2 | Spondin 2 | -0.472 | 0.066 | -7.184 | <0.001 | <0.001 |

* Abbreviations: Std.Err: standard error; Adj.P: adjusted p-value. FC: fold change.

**Table S8**. The top 10 p-values from multivariable linear models that assessed pathway enrichment score change from before to 24 hours after surgery **in the MARBLE cohort**. The linear models were adjusted for the following covariates: CN-105 (all dose groups vs placebo), age, sex (male as the reference group), education years, ApoE-e2 copy number (0, 1, or 2 alleles), ApoE-e4 copy number (0, 1, or 2 alleles), BMI, surgery type (thoracic vs others), ASA physical status classification (1 or 2 as the reference group), and baseline pathway enrichment score. The top panel displays upregulated pathways; the bottom panel displays down-regulated pathways. Beta coefficients are from linear models that assessed the effect of time on the single sample gene set enrichment analysis (ssGSEA) pathway enrichment scores; a positive beta coefficient indicates an increase in the pathway from before to 24 hours after surgery, and *vice versa*.

| **Up-regulated pathways** | **Beta** | **Std.Err** | **t** | **p-value** | **Adj.P** |
| --- | --- | --- | --- | --- | --- |
| negative regulation of smooth muscle cell migration | 84.532 | 10.185 | 8.299 | <0.001 | <0.001 |
| negative regulation of endothelial cell apoptotic process | 101.615 | 12.325 | 8.245 | <0.001 | <0.001 |
| regulation of extrinsic apoptotic signaling pathway via death domain receptors | 94.718 | 13.120 | 7.219 | <0.001 | <0.001 |
| extrinsic apoptotic signaling pathway via death domain receptors | 78.021 | 10.867 | 7.180 | <0.001 | <0.001 |
| smooth muscle cell migration | 46.717 | 7.145 | 6.538 | <0.001 | <0.001 |
| muscle cell migration | 40.831 | 6.534 | 6.250 | <0.001 | <0.001 |
| regulation of muscle system process | 45.909 | 7.608 | 6.034 | <0.001 | <0.001 |
| negative regulation of extrinsic apoptotic signaling pathway via death domain receptors | 87.776 | 15.065 | 5.827 | <0.001 | <0.001 |
| positive regulation of neural precursor cell proliferation | 67.617 | 11.671 | 5.794 | <0.001 | <0.001 |
| regulation of fibrinolysis | 47.596 | 8.313 | 5.725 | <0.001 | <0.001 |
| **Down-regulated pathways** | **Beta** | **Std.Err** | **t** | **p-value** | **Adj.P** |
| liposaccharide metabolic process | -76.872 | 11.187 | -6.871 | <0.001 | <0.001 |
| proteoglycan metabolic process | -56.211 | 9.240 | -6.083 | <0.001 | <0.001 |
| oligosaccharide metabolic process | -57.409 | 9.495 | -6.046 | <0.001 | <0.001 |
| glycoprotein metabolic process | -45.583 | 7.682 | -5.934 | <0.001 | <0.001 |
| sulfur compound metabolic process | -37.941 | 6.490 | -5.846 | <0.001 | <0.001 |
| glycoprotein biosynthetic process | -45.218 | 8.186 | -5.523 | <0.001 | <0.001 |
| proteoglycan biosynthetic process | -51.179 | 9.602 | -5.330 | <0.001 | <0.001 |
| response to chemokine | -79.284 | 15.031 | -5.275 | <0.001 | <0.001 |
| membrane lipid catabolic process | -62.283 | 11.877 | -5.244 | <0.001 | <0.001 |
| negative regulation of leukocyte apoptotic process | -78.409 | 15.015 | -5.222 | <0.001 | <0.001 |

* Abbreviations: Std.Err: standard error; Adj.P: adjusted p-value.

**Table S9**. List of the top 10 p-values from multivariable linear models that assessed the effect of CN-105 treatment (all dose groups vs placebo) on protein intensity changes from before to 24 hours after surgery **in the MARBLE cohort**. The linear models were adjusted for the following covariates: age, sex (male as the reference group), education years, ApoE-e2 copy number (0, 1, or 2 alleles), ApoE-e4 copy number (0, 1, or 2 alleles), BMI, surgery type (thoracic vs others), ASA physical status classification (1 or 2 as the reference group), and baseline protein intensity. Positive Log_2_ Fold Change (Log_2_ FC) values indicate a greater protein level increase among CN-105- vs placebo-treated patients, and *vice versa*.

| **Up-regulated proteins** | | **Log_2_ FC** | **Std.Err** | **t** | **p-value** | **Adj.P** |
| --- | --- | --- | --- | --- | --- | --- |
| SYNE1 | Spectrin Repeat Containing Nuclear Envelope Protein 1 | 0.578 | 0.192 | 3.017 | 0.003 | 0.942 |
| S38AA | Solute Carrier Family 38 Member 10 | 0.120 | 0.041 | 2.912 | 0.004 | 0.942 |
| SMOC2 | SPARC Related Modular Calcium Binding 2 | 0.312 | 0.107 | 2.907 | 0.004 | 0.942 |
| ABCA2 | ATP Binding Cassette Subfamily A Member 2 | 0.110 | 0.040 | 2.767 | 0.007 | 0.942 |
| KAT6B | Lysine Acetyltransferase 6B | 0.588 | 0.214 | 2.743 | 0.007 | 0.942 |
| FUCO2 | Alpha-L-Fucosidase 2 | 0.101 | 0.038 | 2.687 | 0.008 | 0.942 |
| KLK7 | Kallikrein Related Peptidase 7 | 0.246 | 0.096 | 2.553 | 0.012 | 0.967 |
| ADA28 | ADAM Metallopeptidase Domain 28 | 0.140 | 0.055 | 2.547 | 0.012 | 0.967 |
| APOC4 | Apolipoprotein C4 | 0.393 | 0.156 | 2.523 | 0.013 | 0.967 |
| PPIB | Peptidylprolyl Isomerase B | 0.059 | 0.024 | 2.439 | 0.016 | 0.967 |
| **Down-regulated proteins** | | **Log_2_ FC** | **Std.Err** | **t** | **p-value** | **Adj.P** |
| SCUB1 | Signal Peptide, CUB Domain And EGF Like Domain Containing 1 | -0.190 | 0.055 | -3.462 | <0.001 | 0.942 |
| MEGF6 | Multiple EGF Like Domains 6 | -0.363 | 0.109 | -3.334 | 0.001 | 0.942 |
| SRGN | Serglycin | -0.265 | 0.087 | -3.038 | 0.003 | 0.942 |
| FBN1 | Fibrillin 1 | -0.068 | 0.023 | -3.025 | 0.003 | 0.942 |
| SPB9 | Serpin Family B Member 9 | -0.367 | 0.123 | -2.982 | 0.003 | 0.942 |
| MFAP5 | Microfibril Associated Protein 5 | -0.227 | 0.079 | -2.868 | 0.005 | 0.942 |
| DSG2 | Desmoglein 2 | -0.084 | 0.029 | -2.858 | 0.005 | 0.942 |
| MIF | Macrophage Migration Inhibitory Factor | -0.122 | 0.044 | -2.777 | 0.006 | 0.942 |
| AK1A1 | Aldo-Keto Reductase Family 1 Member A1 | -0.306 | 0.110 | -2.772 | 0.006 | 0.942 |
| LDLR | Low Density Lipoprotein Receptor | -0.166 | 0.061 | -2.747 | 0.007 | 0.942 |

* Abbreviations: Std.Err: standard error; Adj.P: adjusted p-value. FC: fold change.

**Table S10.** The top 10 p-values from the multivariable linear models that assessed the effect of CN-105 (all dose groups vs placebo) on pathway enrichment score changes from before to 24 hours after surgery **in the MARBLE cohort**. The linear models were adjusted for the following covariates: age, sex (male as the reference group), education years, ApoE-e2 copy number (0, 1, or 2 alleles), ApoE-e4 copy number (0, 1, or 2 alleles), BMI, surgery type (thoracic vs others), ASA physical status classification (1 or 2 as the reference group), and baseline pathway enrichment score. The top panel displays the pathways that are up-regulated. The bottom panel displays the pathways that are down-regulated. Beta coefficients are from linear models that assessed the effect of CN-105 on the single sample gene set enrichment analysis (ssGSEA) enrichment scores; positive beta coefficients indicate an upregulation of the pathway due to CN-105 (vs placebo) treatment, and *vice versa*.

| **Up-regulated pathways** | **Beta** | **Std.Err** | **t** | **p-value** | **Adj.P** |
| --- | --- | --- | --- | --- | --- |
| leukocyte tethering or rolling | 39.568 | 11.528 | 3.432 | <0.001 | 0.811 |
| hormone transport | 15.314 | 4.925 | 3.109 | 0.002 | 0.915 |
| negative regulation of secretion | 19.645 | 6.429 | 3.056 | 0.003 | 0.915 |
| glycoprotein metabolic process | 17.319 | 6.949 | 2.492 | 0.014 | 0.999 |
| signal release | 13.132 | 5.292 | 2.481 | 0.014 | 0.999 |
| glycoprotein biosynthetic process | 17.525 | 7.366 | 2.379 | 0.019 | 0.999 |
| insulin receptor signaling pathway | 22.504 | 9.483 | 2.373 | 0.019 | 0.999 |
| positive regulation of dna metabolic process | 23.514 | 9.948 | 2.364 | 0.020 | 0.999 |
| n glycan processing | 35.091 | 14.852 | 2.363 | 0.020 | 0.999 |
| proteoglycan biosynthetic process | 20.266 | 8.645 | 2.344 | 0.021 | 0.999 |
| **Down-regulated pathways** | **Beta** | **Std.Err** | **t** | **p-value** | **Adj.P** |
| regulation of muscle contraction | -24.839 | 9.249 | -2.685 | 0.008 | 0.999 |
| regulation of morphogenesis of an epithelium | -20.719 | 7.735 | -2.679 | 0.008 | 0.999 |
| transforming growth factor beta production | -17.526 | 6.581 | -2.663 | 0.009 | 0.999 |
| regulation of striated muscle contraction | -28.941 | 11.084 | -2.611 | 0.010 | 0.999 |
| exocrine system development | -18.568 | 7.112 | -2.611 | 0.010 | 0.999 |
| salivary gland development | -18.850 | 7.523 | -2.506 | 0.013 | 0.999 |
| regulation of animal organ morphogenesis | -16.926 | 6.759 | -2.504 | 0.014 | 0.999 |
| regulation of cardiac muscle contraction | -28.953 | 12.256 | -2.362 | 0.020 | 0.999 |
| regulation of protein dephosphorylation | -30.351 | 12.912 | -2.351 | 0.020 | 0.999 |
| unsaturated fatty acid metabolic process | -26.480 | 11.473 | -2.308 | 0.023 | 0.999 |

* Abbreviations: Std.Err: standard error; Adj.P: adjusted p-value.

Supplemental Figure S2 – volcano plot for univariable effect of CN-105 on preoperative to 24 hr postoperative protein level changes **in the MARBLE cohort**.

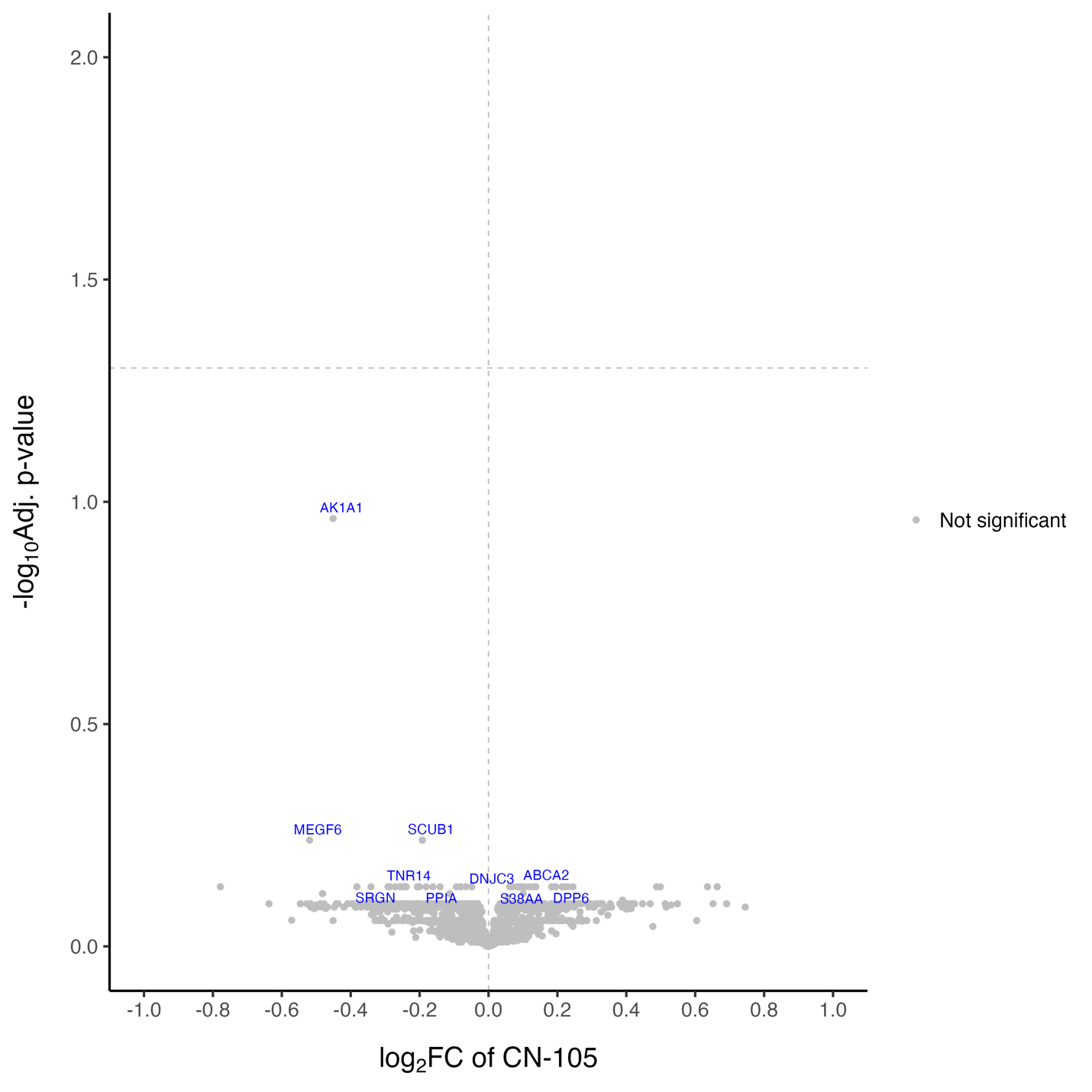

Supplemental Figure **S3** - volcano plot for univariable effect of CN-105 on preop to 24 hr postoperative pathway level changes **in the MARBLE cohort**.

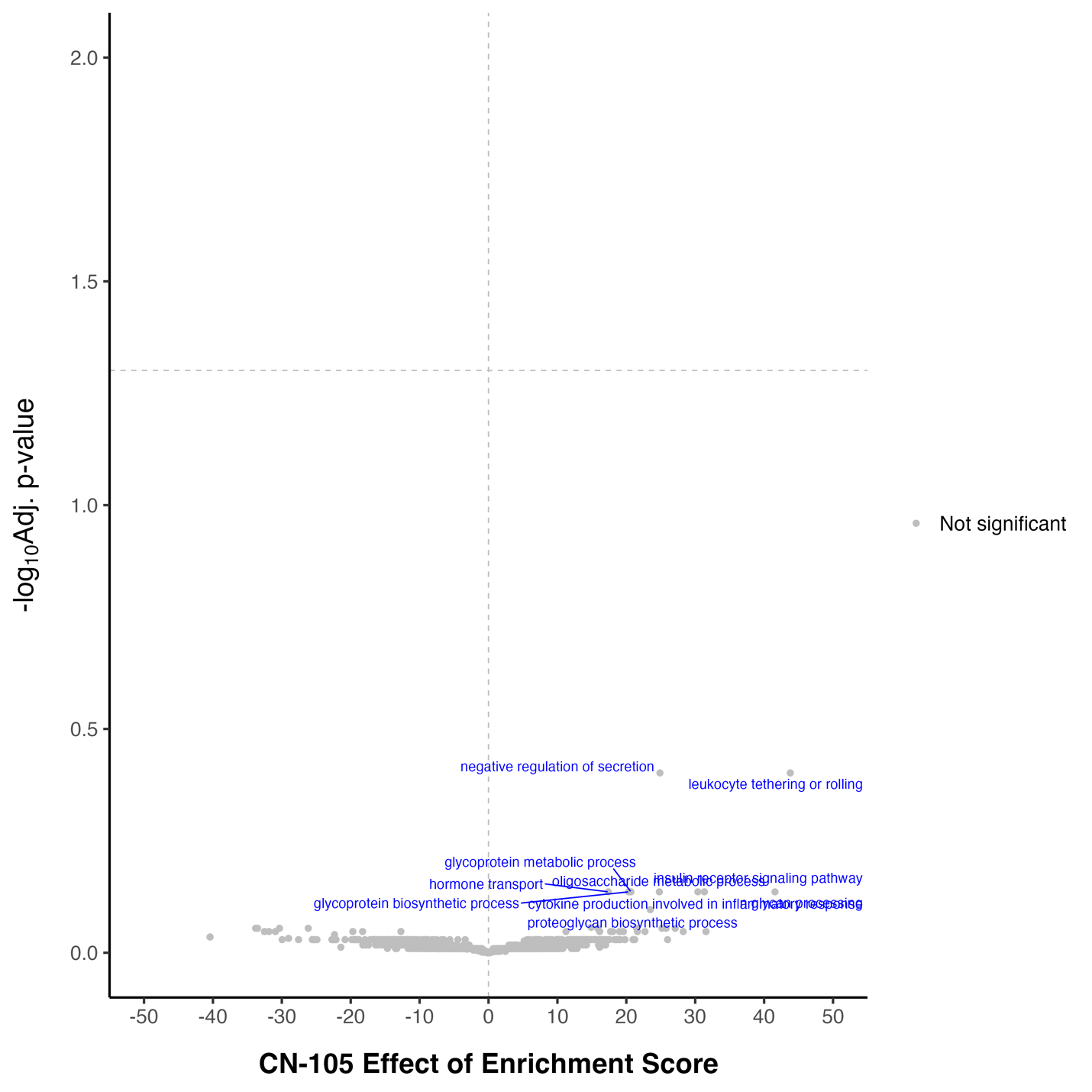

**Section 3: Replication Cohort Description, Demographics, Analysis Strategy.**

**Replication Cohort Description:**

Markers of Alzheimer’s Disease and neuroCognitive Outcomes after Perioperative Care (MADCO-PC) was an observational cohort study, enrolled 140 older surgical patients (age ≥60) undergoing non-cardiac, non-neurologic surgery, and investigated the extent of correlations between postoperative changes in cognitive function and AD-related CSF biomarkers.^7^ Investigating NeuroinflammaTion UnderlyIng postoperative cogniTive dysfunction (INTUIT) was an observational cohort study, enrolled 201 older surgical patients (age ≥60) undergoing non-cardiac, non-neurologic surgery, and investigated the extent to which postoperative changes in CSF inflammatory measures were associated with postoperative changes in cognition, resting state functional MRI brain connectivity and CSF AD-related biomarkers.^8^ Participants in MADCO-PC and INTUIT provided informed consent prior to study participation. CSF proteomic data was available for 96 patients (N=29 from MADCO-PC, N=67 participants from INTUIT) from a separate case/control study of CSF proteomic changes in postoperative delirium, in which patients who developed delirium and those who did not were matched by age, sex, race, surgery group, and van wal raven score (comorbidity burden score).

**Table S11.** Baseline characteristics of the 96 participants from the **Replication cohort**.

|  | N = 96* |
| --- | --- |
| **Age** | 69.9 (5.8) |
| **Sex** |  |
| Male | 63 (65.6%) |
| Female | 33 (34.4%) |
| **Education (years)** | 15.1 (3.3) |
| **Race** |  |
| Caucasian/White | 77 (80.2%) |
| Black or African American | 17 (17.7%) |
| Other | 2 (2.1%) |
| **ApoE-e2 copy number (missing = 1)** | |
| 0 | 82 (85.4%) |
| 1 | 13 (13.5%) |
| 2 | 0 (0%) |
| **ApoE-e4 copy number (missing = 1)** | |
| 0 | 63 (65.6%) |
| 1 | 32 (33.3%) |
| 2 | 0 (0%) |
| **BMI** | 28.3 (4.7) |
| **Surgery type** | |
| Thoracic Surgery | 11 (11.5%) |
| General/Abdominal/Plastic/ENT | 27 (28.1%) |
| Orthopedics Surgery | 23 (24.0%) |
| Urologic Surgery | 35 (36.5%) |
| **ASA physical status classification** | |
| 2 | 27 (28.1%) |
| 3 | 64 (66.7%) |
| 4 | 5 (5.2%) |

*CSF proteomic data was available for 96 patients (N=29 from the MADCO-PC study, N=67 participants from the INTUIT study), from a case/control study of CSF proteomic changes in postoperative delirium, in which patients who did (versus those who did not) develop delirium were matched by age, sex, race, surgery group, and van wal raven score (comorbidity burden score).

**Analysis strategy for protein level changes from before to 24 hours after surgery in the Replication cohort, and comparison to the MARBLE cohort data.**

There were a total of 1887 proteins that overlapped between the two datasets. There were a total of 199 proteins that were uniquely detected in the MARBLE cohort, and 984 proteins that were uniquely detected in the replication cohort. We selected the 881 proteins that showed a significant temporal effect in the MARBLE cohort. Among these 881 proteins, 820 proteins were also detected in the replication cohort. Fitting the univariate linear model that assessed the temporal effect on CSF proteomics change from pre-op to 24-hour post-op to the MARBLE cohort and Replication cohort separately, we derived the log2 fold changes for the 820 proteins that overlap between these two datasets, within each dataset.

We then computed the Spearman’s correlation coefficient for the log2 fold change from pre- to 24-hour post-operative CSF protein levels for each of the 820 proteins in the MARBLE cohort and in Replication cohort, from the univariate linear models on each of these respective cohorts. To compute the Spearman’s correlation coefficient, we extract the log2 fold change of the 820 proteins in the MARBLE cohort (denoted as variable A of length-820) and the log2 fold change of the 820 proteins in the Replication cohort (denoted as variable B of length-820), and then computed the correlation coefficient between these two random variables A and B. The Spearman’s correlation coefficient was 0.861 (95% CI = [0.823, 0.892], p-value<0.001), suggesting a strong alignment of the temporal effect between the two cohorts.

Among these 820 proteins, there were a total of 418 proteins that showed an up-regulation in the MARBLE cohort. This included: (1) 327 proteins that were significantly upregulated after surgery in both MARBLE and the replication cohort; (2) 91 proteins that were significantly upregulated only in the MARBLE cohort. See Supplemental Excel file – Sheet 1 for details.

Among these 820 proteins, there were a total of 402 proteins that showed a down-regulation in the MARBLE cohort. This included: (1) 259 proteins that were significantly downregulated after surgery in both MARBLE and the replication cohort; (2) 143 proteins that were significantly downregulated only in the MARBLE cohort. See Supplemental Excel file – Sheet 2 for details.

***Pathway analysis of Replication cohort, and comparison to MARBLE cohort results***

We performed pathway analysis on the 2871 proteins we detected in the Replication cohort (N= 96 patients) using the Gene Ontology Biological Process database. There were a total of 2375 pathways. We selected the 1001 pathways that showed a significant temporal effect in the MARBLE cohort. Among these 1001 pathways, 932 pathways were also detected in the replication cohort. Fitting the univariate linear model that assessed the temporal effect on CSF pathway change from pre-op to 24-hour post-op to the MARBLE cohort and Replication cohort separately, we derived the enrichment score changes for the 932 pathways that overlap between these two datasets, within each dataset.

We computed the Spearman’s correlation coefficient for the enrichment score (ES) from pre- to 24-hour post-operative timepoint for each of the 932 pathways in the MARBLE cohort and in Replication cohort, from the univariate linear models on each of these respective cohorts. To compute the Spearman’s correlation coefficient, we extracted the ES change of the 932 pathways in the MARBLE cohort (denoted as variable A) and the ES change of the 932 pathways in the Replication cohort (denoted as variable B), and then computed the correlation coefficient between these two random variables A and B. The Spearman’s correlation coefficient was 0.720 (95% CI = [0.680, 0.757], p-value<0.001), suggesting a strong alignment of the temporal effect in pathway enrichment scores between the two cohorts.

Among these 932 pathways, there were a total of 447 pathways that showed an up-regulation in the MARBLE cohort. This included: (1) 261 pathways that were significantly upregulated after surgery in both MARBLE and the replication cohort; (2) 186 pathways that were significantly upregulated only in the MARBLE cohort. See Supplemental Excel file – Sheet 3 for details.

Among these 932 pathways, there were a total of 485 pathways that showed a down-regulation in the MARBLE cohort. This included: (1) 302 pathways that were significantly downregulated after surgery in both MARBLE and the replication cohort; (2) 183 pathways that were significantly downregulated only in the MARBLE cohort. See Supplemental Excel file – Sheet 4 for details.

***Pathway analysis of proteins that showed a significant time effect in both datasets, in the MARBLE dataset.***

We selected 348 proteins that showed a significant time effect in both datasets (up-regulated: 227; down-regulated: 121 proteins), and then performed an ssGSEA pathway analysis on these protein level changes from the preoperative to 24 hour postoperative time point in the MARBLE dataset. Among a total of 448 pathways, there were 359 pathways that showed a significant time effect in the MARBLE cohort. See Supplemental Excel file – Sheet 5 for down-regulated pathways. See Supplemental Excel file – Sheet 6 for up-regulated pathways.

We also performed the ssGSEA pathway analysis on these 348 protein level changes from the preoperative to 24 hour postoperative time point in the Replication dataset. Among a total of 448 pathways, there were 310 pathways that showed a significant time effect. See Supplemental Excel file – Sheet 7 for down-regulated pathways. See Supplemental Excel file – Sheet 8 for up-regulated pathways.
